# Memory B cell proliferation drives differences in neutralising responses between ChAdOx1 and BNT162b2 SARS-CoV-2 vaccines

**DOI:** 10.1101/2024.07.11.24310221

**Authors:** David Hodgson, Yi Liu, Louise Carolan, Siddhartha Mahanty, Kanta Subbarao, Sheena G. Sullivan, Annette Fox, Adam Kucharski

## Abstract

Vaccination against COVID-19 has been pivotal in reducing the global burden of the disease. However, Phase III trial results and observational studies underscore differences in efficacy across vaccine technologies and dosing regimens. Notably, mRNA vaccines have exhibited superior effectiveness compared to Adenovirus (AdV) vaccines, especially with extended dosing intervals. Using in-host mechanistic modelling, this study elucidates these variations and unravels the biological mechanisms shaping the immune responses at the cellular level. We used data on the change in memory B cells, plasmablasts, and antibody titres after the second dose of a COVID-19 vaccine for Australian healthcare workers. Alongside this dataset, we constructed a kinetic model of humoral immunity which jointly captured the dynamics of multiple immune markers, and integrated hierarchical effects into this kinetics model, including age, dosing schedule, and vaccine type. Our analysis estimated that mRNA vaccines induced 2.1 times higher memory B cell proliferation than AdV vaccines after adjusting for age, interval between doses and priming dose. Additionally, extending the duration between the second vaccine dose and priming dose beyond 28 days boosted neutralising antibody production per plasmablast concentration by 30%. We also found that antibody responses after the second dose were more persistent when mRNA vaccines were used over AdV vaccines and for longer dosing regimens. Reconstructing in-host kinetics in response to vaccination could help optimise vaccine dosing regimens, improve vaccine efficacy in different population groups, and inform the design of future vaccines for enhanced protection against emerging pathogens.

**SIGNIFICANCE STATEMENT:** There are differences in vaccine efficacy across different SARS-CoV-2 vaccine technologies and dosing regimens. Using an in-host mechanistic model that describes antibody production fitting to in-host immune markers, we found that mRNA vaccines are twice as effective at stimulating memory B cell proliferation when compared to AdVs vaccines and that a longer time between the second vaccine dose and priming dose increases the neutralising antibody production per plasmablast concentration. These findings disentangle the effect of vaccine type and time since the priming dose, aiding in the understanding of immune responses to SARS-CoV-2 vaccination.

## INTRODUCTION

Vaccination has been a crucial tool in the global reduction of the COVID-19 burden since the approval of early vaccine candidates in December 2020. The earliest vaccines to be approved included an adenoviral-vectored vaccine (i.e. ChAdOx1) and mRNA vaccines (i.e. BNT162b2 and mRNA-1273). However, as Phase III trial results and observational studies emerged, variation was observed in the estimated efficacy and effectiveness of different vaccine types and their corresponding dosing schedules.(1) For example, the efficacy of BNT162b2 against symptomatic COVID-19 after two doses given three weeks apart was initially reported at 95.0% (95% CI 90.3–97.6) for a median follow-up of two months(2) and 91.3% (95% CI 89.0–93.2) after at least 6 months of follow up.(3) The efficacy of ChAdOx1 against symptomatic COVID-19 after two doses given ≤12 weeks apart was initially reported at 62.1% (95% CI 41.0–75.7) after a 53–90 day follow-up.(4). Consistent with these findings, observational studies have found higher effectiveness of the BNT162b2 vaccine compared to the ChAdOx1 vaccine at preventing symptomatic COVID-19.(5) Moreover, the dosing schedule has been shown to influence the vaccine effectiveness of both products, with longer dosing schedules (time between first and second dose) seeing higher effectiveness values than shorter dosing schedules. Specifically, for ChAdOx1, vaccine efficacy was 81·3% [95% CI 60·3–91·2] with a dosing schedule ≥12 weeks and 55·1% [33·0–69·9] at <6 weeks.(4) For BNT162b2, lower risks of symptomatic SARS-CoV-2 infection have been observed when the dosing schedule was extended from 17–25 days to 26–42 days(6)

Despite these heterogeneous observations in efficacy, a comprehensive understanding of the immunological processes underlying the effects of vaccine type and dosing schedule on vaccine efficacy remains elusive. Both ChAdOx1 and BNT162b2 elicit robust cellular responses and cross-reactive neutralising antibodies against different SARS-CoV-2 variants, promoting the persistence and maturation of memory B cells (MBC) over time, and contributing to durable immunity.(7, 8) However, there are also notable differences in the immunological responses. For example, ChAdOx1 triggers robust T cell and antibody responses, particularly generating IgG and IgM antibodies along with Th1 cytokines such as IL-2, TNF-α, and INF-γ.(9, 10) Whereas, BNT162b2 initiates potent B cell responses and antibody secretion, particularly of IgA and IgG, usually at much higher levels than responses to the ChAdOx1 vaccine.(10) When considering the dosing schedule, a longer dose schedule for BNT162b2 resulted in higher neutralizing antibody titres, whilst maintaining comparable T cell responses.(11, 12) Assessing variations in the immunological response to vaccination presents challenges, as the schedules differed between ChAdOx1 and BNT162b2 vaccines upon deployment, complicating the disentanglement of the impact of vaccine type and dosing schedule on immunological kinetics.

Analysis of in-host immunological responses can be used to understand the dynamic behaviour of the factors driving these responses, formalising correlates between immune markers, and allowing identification of key factors that influence the timing and magnitude of immune responses. Such analysis typically adopts a phenomenological approach, aiming to formalise correlations between observed phenomena through a purely statistical approach rather than explicitly elucidating underlying biological mechanisms.(13, 14) Mechanistic in-host models, on the other hand, specify the detailed biological processes driving immune responses, allowing deeper insights into the causal effects of antibody production and immune cell kinetics.(13) However, a significant challenge with mechanistic models is their susceptibility to identifiability issues; where due to limited or noisy data, multiple sets of parameters can produce the same observed outcomes, making it difficult to determine the true underlying biological mechanisms.(15) Establishing a mechanistic model capable of disentangling key processes can therefore enable a more accurate comparison of immunogenicity between vaccine types.

In this study, we employ an in-host mechanistic model to reconstruct the unobserved kinetics of SARS-CoV-2 immune markers and consider the influence of host factors on driving immunological heterogeneity (Figure 1). Specifically, our investigation aims to elucidate mechanistic explanations for differences in vaccine-neutralising activity and MBC kinetics in response to second-dose vaccination against COVID-19. Our in-host model incorporates two sources of antibody production: plasmablasts and plasma cells, both stemming from vaccine-induced differentiation of MBC. By fitting our model to multiple biomarkers, including concentrations of MBC and plasmablasts, as well as surrogate virus neutralisation test (sVNT) titers against ancestral SARS-CoV-2 strains, we delineate the distinct immunogenicity (characterised by the rate of MBC proliferation) and antibody affinity (measured by neutralising sVNT per MBC) profiles associated with ChAdOx1 and BNT162b2 vaccine types. Further, we validate this mechanistic model by predicting antibody kinetics on unseen validation data and show that the predictions remain accurate providing that baseline immune information can be measured for each individual. Finally, we investigate the influence of host factors, such as the time elapsed since initial vaccination, as well as demographic characteristics such as age, on the in-host kinetics of these immunological responses

**Figure 1.**
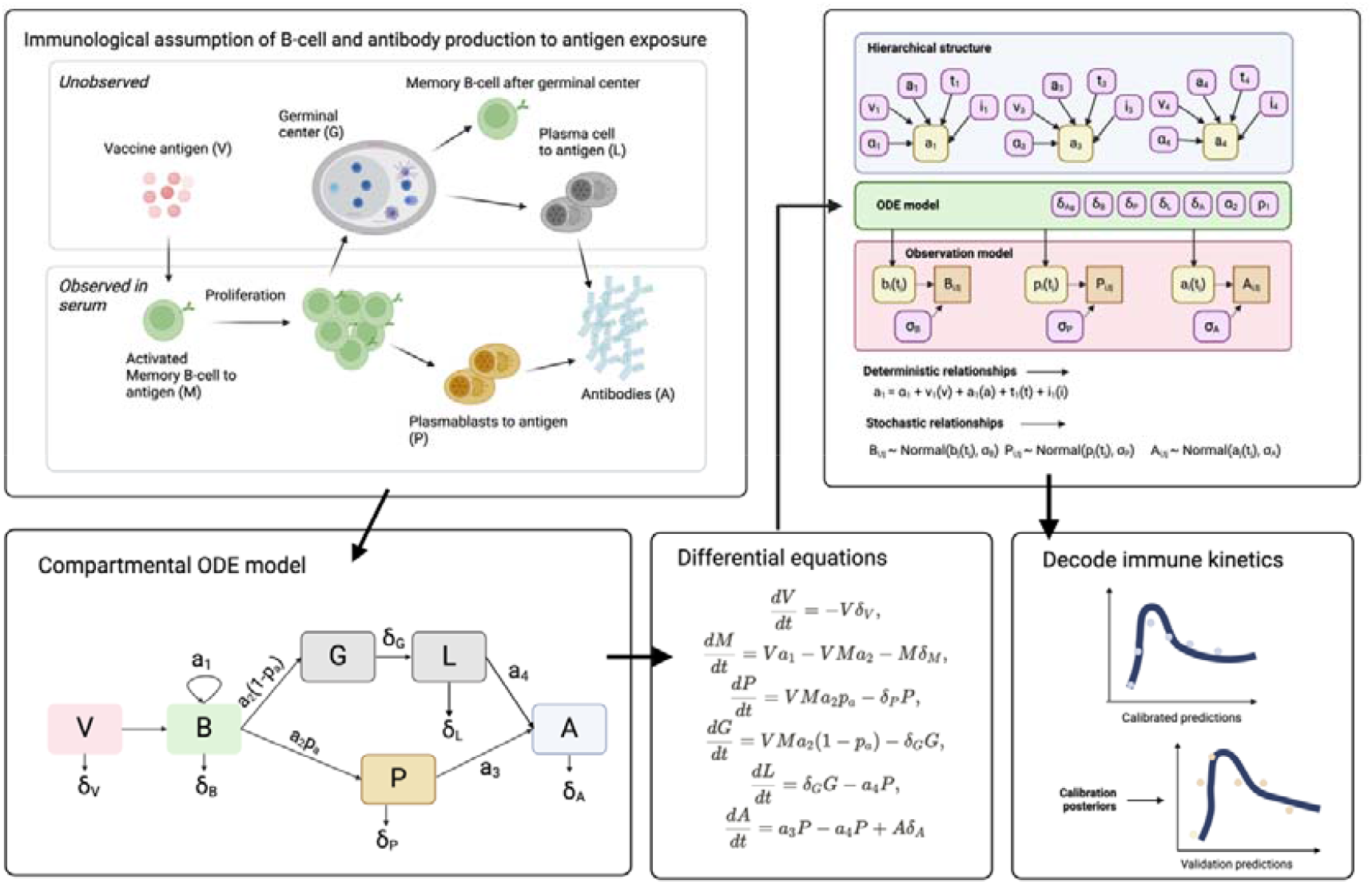
Illustration of the link between the immunological assumptions, dynamic model and the hierarchical effects on the parameters of interest.

## RESULTS

### Model performance on calibration and validation data

We fitted the model to two different antigens expressed on the MBC and plasmablasts: the ancestral spike antigen and the receptor binding domain (RBD) antigen. For each model, we fit to the same antibody data, measured by sVNT assay, which measures antibodies that inhibit RBD binding to ACE2. The sampled posterior distributions for both models were effectively explored, with all parameters demonstrating convergence and the chains showing good mixing (Figures S1–2). After calibrating the two models to the calibration dataset, we found that immune trajectories could reproduce the observed dynamics in the data (Figure 2A). We calculated the Continuous Ranked Probability Score (CRPS) to assess the goodness-of-fit between the two models and found that for all three immune markers (MBC, plasmablasts and sVNT) similar scores were achieved for both models (Figure 2B). We then evaluated the predictive accuracy of the fitted model by using baseline estimates from the validation dataset (immune markers before vaccination) to predict the trajectories of each biomarker and then assessed the fit using the CRPS. We found that the model predictions to the validation dataset were reflective of the data and that both the spike and RBD models provided similar fits in terms of CRPS (Figure 2C-D). From this, we cannot conclude that either the spike or RBD model is better at describing sVNT in each individual. The individual-level fits for each time point, and the resulting individual-level trajectories for each model are provided in Figures S3–6. The posterior distributions for the fitted distributions of the hierarchical and decay parameters are also provided in Figures S7–10.

**Figure 2.**
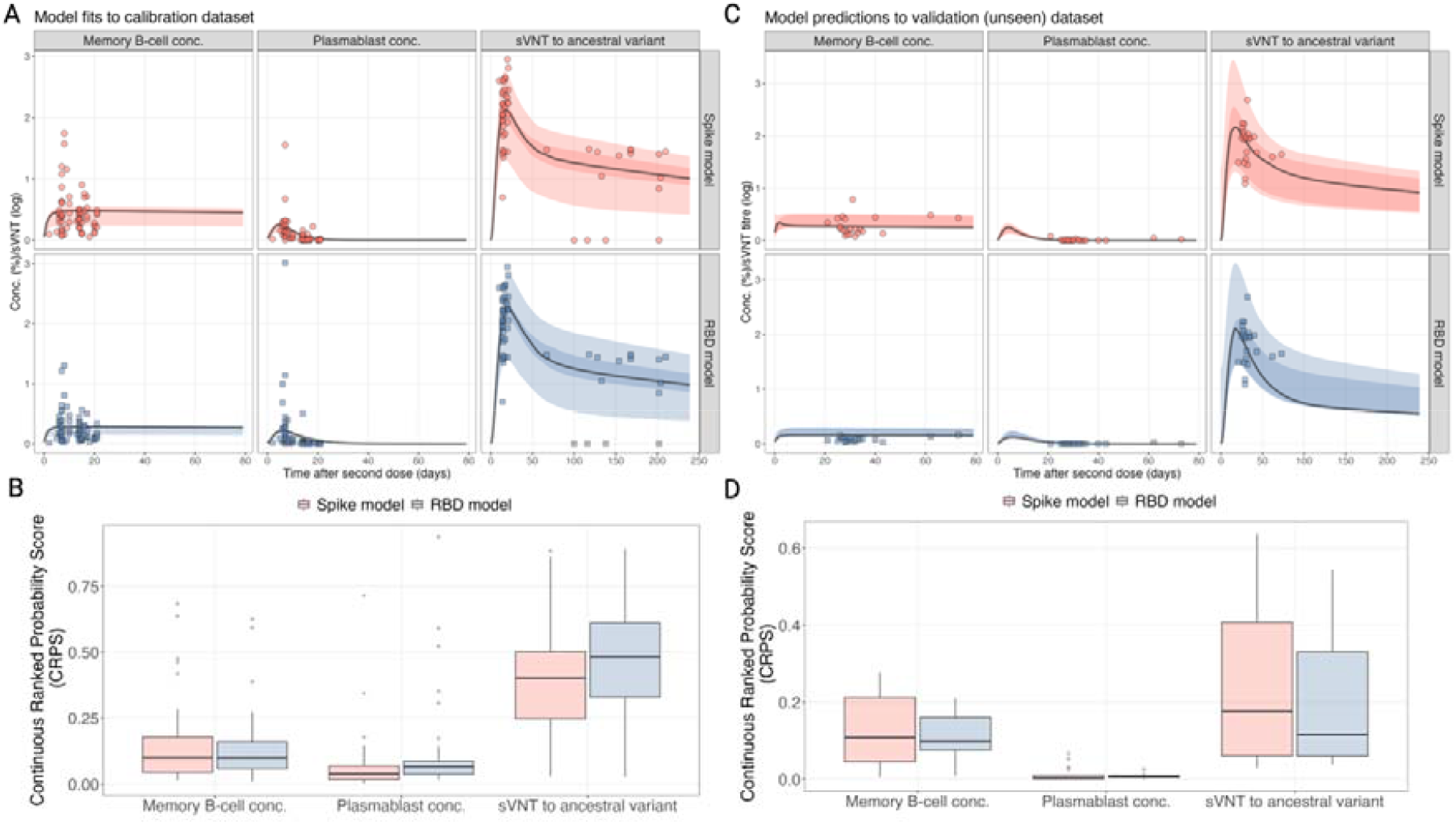
Comparison of the fitted models and raw data from the calibration dataset (A, B) and the validation dataset (C, D). (A, C) Comparison of model posterior predictive trajectories, fitted to each antigen (rows), for the biomarker type (columns) for time post-vaccination (x-axis). (B, D) The distribution of the CRPS for both models fits for each biomarker for the calibration dataset.

### Drivers of MBC proliferation and antibody production

By combining multiple biomarkers with a dynamic model, we were able to reconstruct individual-level kinetics of MBC frequencies, plasmablast frequencies and sVNT as well as estimate overall average population kinetics for these markers. In the process, we were able to estimate three key mechanistic processes underlying the observed kinetics: the rate of MBC proliferation (a1), the antibody affinity of plasmablasts (a3), and the antibody affinity of plasma cells (a4). For the spike model, we estimated that the BNT162b2 vaccine induced substantially more MBC proliferation when compared to ChAdOx1, with ancestral spike reactive MBC concentrations (of total MBC) increasing by 0.44% (95% PPI 0.28–0.58) vs. 0.21% (95% PPI 0.11–0.34) per day per vaccine unit (Figure 3A). Lower rates of MBC proliferation were seen in the RBD model, with concentration increasing by 0.26 (95% PPI 0.16–0.25) and 0.13 (95% PPI 0.06–0.22) per day per vaccine unit for BNT162b2 and ChAdOx1 respectively. We also estimated that age and the dosing schedule had little impact on the rate of B cell proliferation after adjusting for vaccine type, with no notable trend between age group levels (Figure 3B and Figure S11A).

**Figure 3.**
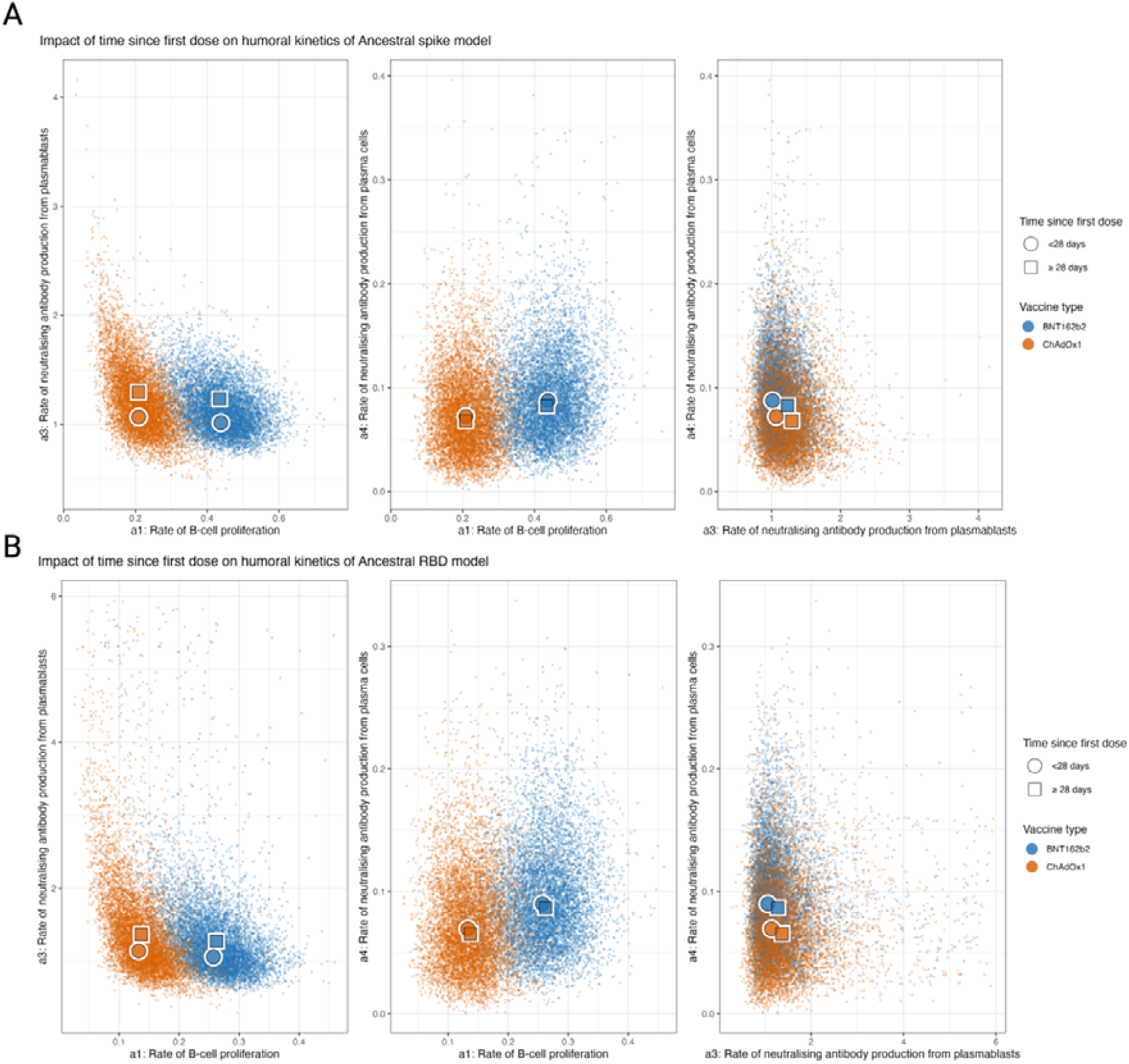
Posterior predictive distributions of the parameters driving the in-host mechanistic model according to key covariates for ancestral spike and RBD stratified by time since the first dose. (A) Posterior predictive distributions for the impact of time since the first dose on the rate of MBC proliferation (a1), the rate of sVNT antibody production from plasmablasts (a3), the rate of sVNT antibody production from plasma cells (a4). (B) Posterior predictive distributions for the age on a1, a3, and a4.

However, for the rate of production of sVNT antibodies per plasmablast, we found that dose interval had a notable impact, with those vaccinated <28 days prior seeing lower rates of sVNT antibodies produced per plasmablast concentration per day (1.08 (95% PPI 0.74– 1.66) when compared to those vaccinated ≥28 days prior) (1.30 (95% PPI 0.90–2.94)) for the spike model. Similar estimates were also found for the RBD model with rates of antibody production of 1.26 (95% PPI 0.67–2.96) for <28 days compared to 1.48 (95% PPI 0.80–3.26) for RBD). We also found that the rate of production of neutralising antibodies per plasma cell remains similar across vaccine type, time since last dose and age group. These observations remained consistent for ancestral RBD.

### Mechanistic predictions of antibody kinetics

By inferring underlying in-host kinetics, we could also estimate the temporal variation in the origin of antibody production following vaccination with a second dose (Figure 4). For ancestral spike, after vaccination with ChAdOx1, our findings reveal a peak in antibody titres 16 days post-vaccination with a log2 sVNT of 1.29 (95% PPI 0.97–1.66). These titres then undergo a fast period of waning until around 50 days and then have a slower period of waning driven by antibody production from plasma cells, gradually declining to an sVNT of 0.48 (95% PPI 0.21–0.74) by day 250 post-vaccination. For the BNT162b2 vaccine, we find a peak antibody titre at day 17 post-vaccination with a log2 sVNT of 2.31 (95% PPI 2.13– 2.50), which then wane to an sVNT of 1.09 (95% PPI 0.80–1.40) at day 250 post-vaccination. The time at which antibodies produced from plasmablasts dominate the sVNT response (transition time in Figure 4A) is 54 and 51 days for ChAdOx1 and BNT162b2 vaccines, respectively. Similar trends are observed for the ancestral RBD model and are summarised in Figure S12A.

**Figure 4.**
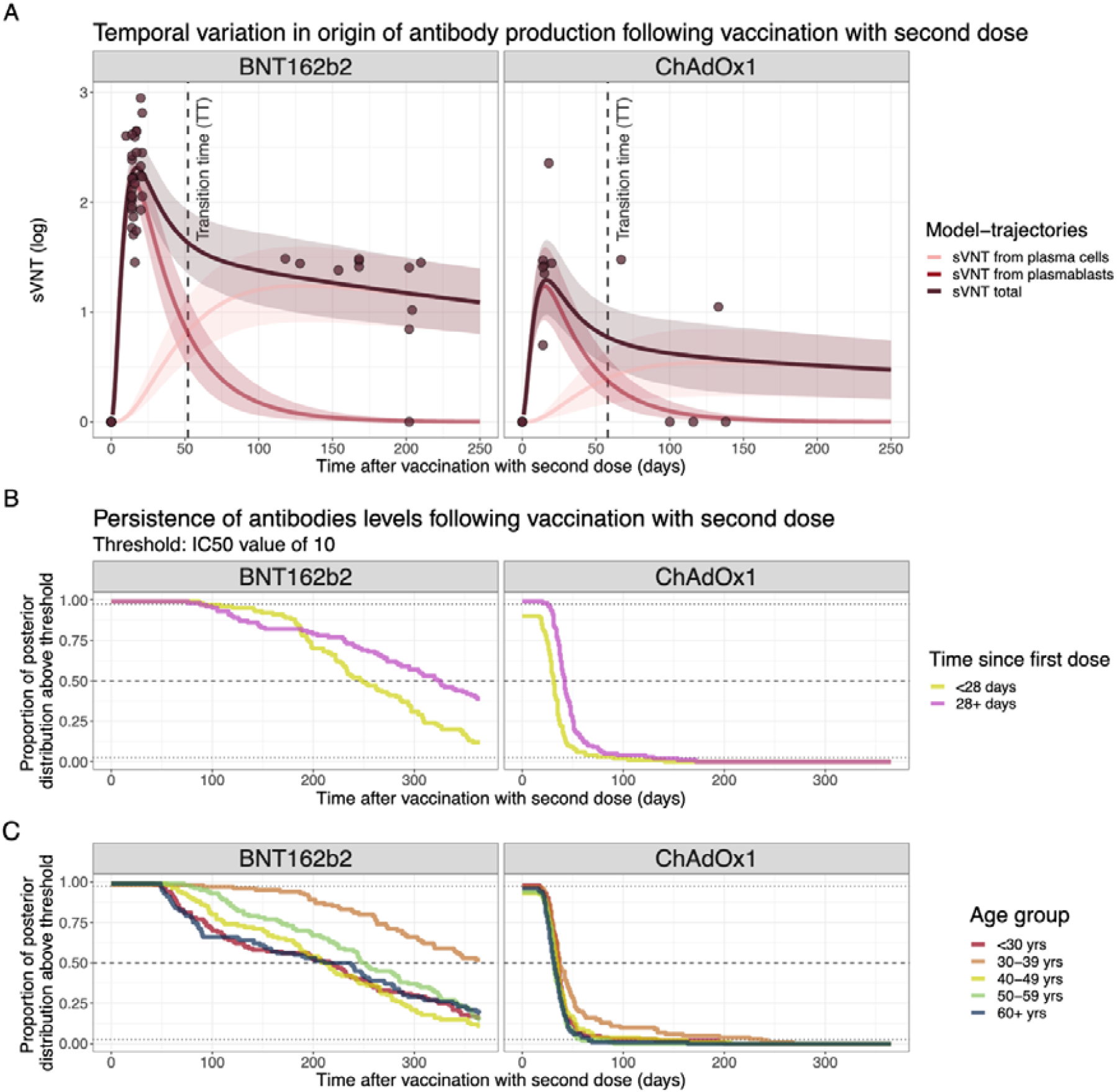
A) Source of sVNT antibodies by days post-vaccination for ancestral RBD. Line and ribbons show the mean and 95% posterior predictive interval (PPI) and dots represent the sVNT from data. (B-–C) Complementary CDF of the marginal posterior distributions for the time antibody titres remain above an IC50 threshold of 10 stratified by vaccine type and B) time since the last dose or C) age group.

We also estimated the duration after vaccination that an individual sVNT IC50 titre remained above 10 by assessing the proportion of the posterior distribution that is above this threshold as a function of time since vaccination. We estimate that the median duration of time that sVNT IC50 titres exceeded 10 was 290 (95% PPI 105–365) days and 34 (95% PPI 0–96) days for BNT162b2 and ChAdOx1, respectively. When stratified by time since first dose, we estimated that individuals who were vaccinated <28 days since their first dose maintained IC50 titres in excess of 10 for shorter periods compared to those with longer time between doses, i.e. 30 (95% PPI 0–90) and 50 (95% PPI 25–130) days for ChAdOx1 respectively, and 247 (95% PPI 91–365) and 33 (95% PPI 88–365) days for BNT162b2 vaccine respectively (Figure 4B and Figure S12B). No strong trend was seen with antibody persistence with increasing age (Figure 4C and Figure 12C).

## DISCUSSION

Using an in-host mechanistic model, we reconstructed the mechanisms driving the production of antibodies from two MBC sources in response to a second dose of ChAdOx1 and BNT162b2 SARS-CoV-2 vaccine. Our estimation of the MBC proliferation rate and antibody production revealed significantly higher responses among BNT162b2 vaccinees compared to ChAdOx1 vaccinees, but age and dosing schedule showed minimal impact on MBC proliferation rates. Instead, we found that dosing interval affected the rate of neutralising antibody production per plasmablast, with lower rates observed for those receiving their second dose within 28 days of their first dose compared to those who received their second dose after 28 days. In-host kinetic modelling made it possible to reconstruct peak antibody titres and duration of antibody persistence, indicating longer periods of protection with BNT162b2 when compared to ChAdOx1, and longer periods of protection for individuals vaccinated more than 28 days since their first dose. These findings suggest that vaccine implementation efforts should consider the intricate interplay between dosing schedule and immune kinetics to optimise vaccine efficacy and durability.

## DISCUSSION

Using an in-host mechanistic model, we reconstructed the mechanisms driving the production of antibodies from two MBC sources in response to a second dose of ChAdOx1 and BNT162b2 SARS-CoV-2 vaccine. Our estimation of the MBC proliferation rate and antibody production revealed significantly higher responses among BNT162b2 vaccinees compared to ChAdOx1 vaccinees, but age and dosing schedule showed minimal impact on MBC proliferation rates. Instead, we found that dosing interval affected the rate of neutralising antibody production per plasmablast, with lower rates observed for those receiving their second dose within 28 days of their first dose compared to those who received their second dose after 28 days. In-host kinetic modelling made it possible to reconstruct peak antibody titres and duration of antibody persistence, indicating longer periods of protection with BNT162b2 when compared to ChAdOx1, and longer periods of protection for individuals vaccinated more than 28 days since their first dose. These findings suggest that vaccine implementation efforts should consider the intricate interplay between dosing schedule and immune kinetics to optimise vaccine efficacy and durability.

Serological analysis of antibody responses of cohorts with homologous two-dose vaccination schedules shows that those with BNT162b2 have higher levels of sVNT to ancestral variants compared to ChAdOx1.(10) We find this difference is driven by 2.1 times higher rates of MBC proliferation caused by BNT162b2, compared to ChAdOx1, and not by differences in antibody affinity per antibody-secreting cell. This could indicate that mRNA vaccines are better at stimulating B cells, potentially due to the amount of antigen produced or to co-stimulatory and inflammatory signals.(16) We present findings from our study of a healthy cohort aged 18–60, revealing no correlation between the in-host kinetics of vaccine-induced humoral responses and age. This observation aligns with broader immunological insights, which show that age-related variations in the immune response to vaccination are exhibited primarily in children and adults aged 65 years and older.(17–20)

Our study also revealed that delaying the time until the second dose led to higher affinity antibodies from plasmablasts, causing more robust post-vaccination sVNT. A longer dosing schedule has previously been shown to lead to higher sVNT and a greater magnitude of mature MBC post-vaccination.(11, 12, 21) This is likely driven by increased affinity maturation in B cells, which continues up to 6 months after vaccination.(22, 23). Vaccinating individuals whilst affinity maturation is ongoing could lead to sub-optimal antibody repertoires being proliferated in-host and thus lead to reduced antibody affinity compared to those with longer dosing schedules.(24) Whilst the dosing schedule influences antibody affinity, we found it had little impact on the rate of proliferation of B cells or the antibody affinity of plasma cells. Given its notable influence on antibody affinity, public health strategies should consider the importance of the dosing schedule when offering boosting campaigns to maximize vaccine-induced protection, particularly for those where strong and lasting immunity is critical.

We were able to mechanically describe the kinetics of antibody responses following a second dose of SARS-CoV-2 vaccination. Peak antibody levels were observed at 17 days post-vaccination (consistent with previous modelling estimates of 15 days (25)) and plasma cell responses were found to dominate the sVNT response after approximately 50 days. Our model also estimated that plasmablast concentration peaked at five days after a second vaccine dose and returned to baseline at 19 days. These dynamics are corroborated by observational studies, such as Pape et al. (2021),(26) which reported a peak in spike-binding plasmablasts to second dose mRNA vaccines at 5 days post-vaccination, returning close to baseline by day 11. Similarly, Turner et al. (2021),(27) found plasmablast responses to vaccine doses returning to baseline by 2 weeks. In addition, our hierarchical regression analysis allowed us to consider marginal posterior distributions and account for potential confounding with dosing schedules, estimating the relative impact of adenovirus (AdV) vaccines if dosing schedules were shorter. By estimating the antibody persistence if the ChAdOx1 dosing schedule was less than 28 days, a counterfactual outcome, we can disentangle the effect of vaccine type and time since the priming dose, aiding in the understanding of immune responses to SARS-CoV-2 vaccination

Our study has some limitations. Firstly, the sample size was relatively small, comprising only 41 participants in the calibration dataset and 22 in the validation dataset, and both cohorts remained infection naive throughout, making it difficult to generalise these results to other populations who may have had prior infections, which are known to change immune kinetics compared to those who are infection naive.(28) Additionally, our measured observations of immune markers were confined to peripheral blood, potentially overlooking critical immune dynamics within lymphoid organs which may influence antibody kinetics. Further, this study also did not account for Helper T cell interactions, which play a crucial role in regulating the memory immune response. Finally, there are host factors not included in this analysis which could have influenced in-host immune heterogeneity, including age, genetic polymorphism, epigenetic factors and variations in cellular immunity.(29, 30)

Our study highlights the importance of understanding in-host immunological kinetics in future vaccine development instead of relying just on Phase III endpoints. Understanding how age, vaccine type and dose schedule impact immune kinetics allows for the customisation of vaccination strategies tailored to different demographic groups, optimising protection across diverse populations. Furthermore, our findings underscore the importance of ongoing monitoring and surveillance post-vaccination to assess the persistence of immune responses. By integrating real-time data on immune kinetics into vaccine development and deployment strategies, stakeholders can make informed decisions regarding doses, vaccine updates to address emerging variants and allocation of resources in response to developing public health needs. These could be particularly useful in emergency contexts such as the Coalition for Epidemic Preparedness Innovations (CEPI) 100-day mission, which aims to develop and deploy vaccines in lower and middle-income countries in a very short timeframe.(31)

In this study we reconstructed unobserved immunological kinetics and accounted for host factors which vary between individuals. This model extends previous humoral kinetics frameworks by fitting to multiple humoral biomarkers and incorporating hierarchical effects. We have shown that this framework can provide valuable insights into the mechanisms underlying vaccine-induced immune responses and aid in the development of more effective vaccination strategies and the impact of dosing schedules. In turn, a better understanding of in-host immunological kinetics in response to vaccination could help optimise vaccine dosing regimens to maximise vaccine efficacy in different population groups and inform the design of future vaccines for enhanced protection against other emerging pathogens.

## MATERIALS AND METHODS

### Study design of calibration dataset

In April 2020, a prospective, open cohort study (ClinicalTrials.gov Identifier: NCT05110911) was established to investigate influenza vaccine immunogenicity among Health Care Workers (HCWs) at six health services across Australia (Alfred Hospital, Melbourne; Children’s Hospital Westmead, Sydney; John Hunter Hospital, Newcastle; Perth Children’s Hospital, Perth; Queensland Children’s Hospital, Bisbane; and the Women’s and Children’s Hospital, Adelaide). Commencing April 2021, the study pivoted to enable follow-up of COVID-19 vaccination. HCWs, including medical, nursing, and allied health staff, students and volunteers aged 18Y to 60Y, were recruited at each hospital’s staff influenza vaccination clinic or responded to recruitment advertising. Those on immunosuppressive treatment (including systemic corticosteroids) within the past 6 months, and contraindicated for vaccination were excluded. Enrolled participants provided a 9ml blood sample for serum collection prior to a first dose and ∼14 days after their second dose of COVID-19 vaccine (suggested range 10-21 days) and at the end of the year. Pre-vaccination blood samples were collected from participants upon enrolment for participants newly enrolled in 2021, or samples collected in late 2020 were used for participants already enrolled in the influenza vaccination cohort in 2020. End-of-year sera were collected October through November 2021. A subset of 41 participants provided additional blood samples for peripheral blood mononuclear cell (PBMC) recovery on day 0 if enrolled prior to receiving their first vaccine dose and ∼ 7 and 14 days after vaccination.

The study protocol and protocol addendums for follow-up of COVID-19 vaccinations and SARS-CoV-2 infections were approved by The Royal Melbourne Hospital Human Research Ethics Committee (HREC/54245/MH-2019). LSHTM Observational Research Ethics Committee of London School of Hygiene and Tropical Medicine gave ethical approval for the use of this data for analysis (ref 22631).

### Study design of validation dataset

Samples for the validation dataset (n=22) were collected in a prospective observational study of immune responses to COVID-19 vaccines conducted at the Royal Melbourne Hospital and the Peter Doherty Institute for Infection and Immunity in Melbourne from June 2020 to December, 2022, funded by the National Institutes of Health, Bethesda, MD (the DISCOVER-HCP-BOOSTER study, HHSN272201400005C). In this study in health care providers (including clinical and allied health staff at the two institutions), after informed consent was obtained, blood samples (∼50 ml/sample) were collected before the first dose of either ChAdOx1 (n=15) or BNT162b2 vaccines (n=10), then 3-4 weeks after the first dose, just before the second dose (which occurred 3 weeks after the first dose for BNT162b2 recipients) and 11 weeks after the first dose for ChAdOx1 recipients) and 2-4 weeks after the second dose for all vaccine recipients. PBMC were isolated within 6 hours of collection and cryopreserved in liquid nitrogen until analysis.

The study protocol and all samples collected in the DISCOVER-HCP-BOOSTER study were approved after review by the RMH Human Research and Ethics Committee (HREC/63096/MH-2020).

### Surrogate Virus Neutralization Test (sVNT) assay

The SARS-CoV-2 sVNT assay described by Tan et al(32) was adapted to utilize commercially available SARS-CoV-2 spike receptor binding domain (RBD) protein (SinoBiological, 40592-V27H-B) representative of the ancestral strain (YP_009724390.1). Sera were serially diluted 3-fold from 1:10 to 1:21870 for testing. GraphPad Prism version 9.5.1 for Windows (GraphPad Software, California USA) was used to fit sigmoidal curves of OD450 values against log10 serum dilutions and to interpolate 50% inhibition titres. Sera that had no detectable inhibition at the lowest dilution were assigned a value of 1. Full details are provided in a prior publication.(8)

### SARS-CoV-2 spike- and RBD-reactive B cell analysis

PBMCs were recovered using Lymphoprep (STEMCELL Technologies, Vancouver, Canada) and LEUCOSEP tubes (Greiner); cryopreserved in FCS containing 10% DMSO; and thawed into RPMI containing DENARASE (cLEcta, Leipzig Germany). Biotinylated spike and RBD were labelled with Streptavidin-fluorochromes (SA-F). PBMCs were incubated with fluorescent-labeled recombinant spike and RBD proteins and with a cocktail of mAbs to detect activated MBC. Full details are provided in a prior publication.(8)

### Dynamic modelling of in-host kinetics to vaccination

Our aim was to develop a mechanistic model of antibody production to gain a deeper understanding of the relationship between antibody-secreting cells, such as plasma cells and plasmablasts, and the changes in serum antibody concentrations over time following a second dose of SARS-CoV-2 vaccination. Our objectives were threefold: i) Establish a mechanistic model of antibody production in response to COVID-19 vaccination, ii) Determine the host factors driving immune heterogeneity to vaccination and iii) Determine the temporal variation in the origin of antibody production over time.

For the measured biomarkers, we devised an in-host mathematical model of humoral immunity to assess the kinetics of antibody production for each vaccine type. We assume that the vaccine antigen stimulates the proliferation of memory B cells. After this stimulation, these cells differentiate along one of two pathways. They can either become short-lived plasmablasts, which secrete an initial burst of antibodies in response to vaccination, or they can migrate to the germinal centre. In the germinal centre, they remain for approximately two weeks before differentiating into long-lived plasma cells, which also secrete antibodies. Incorporating hierarchical effects into the model will help us assess the impact of host factors on the kinetics of memory B cells and antibodies. Identifying how previous vaccination history—considering vaccine type and timing—affects immune responses will inform improvements in vaccine formulations. Understanding the variability in immune responses due to host factors will enable personalised vaccination strategies to enhance overall vaccine effectiveness. Finally, by analysing the kinetics within the fitted dynamic models, we aim to understand the timeline of antibody production, including the transition from short-term to long-term immunity and the roles of different immune cells and organs over time. This temporal analysis will help predict the duration of immunity provided by the vaccine, thereby informing public health policies on optimal vaccination schedules and the frequency of booster doses.

### Likelihood Function

The likelihood function assumes that measured biomarkers are subject to normal distribution errors. The combined likelihood relates the data for MBC, plasmablasts, and antibodies to their model-predicted quantities. The details of the likelihood equations are given in the Supplementary Information. Priors for the dynamic system parameters are based on immunological observations. For instance, the decay rates of various cell types and antibodies are assigned priors reflecting their known biological behaviour. Non-informative and weakly informative priors are used for other parameters to ensure flexibility while maintaining biological plausibility. A full list of priors and their derivations is given in the Supplementary Information.

### Implementation

The model is implemented using Hamiltonian Monte Carlo (HMC) via Stan. The ODEs are solved at each Markov chain step using the Runge-Kutta method. The analysis is conducted on two datasets: one using memory B cells and plasmablasts for ancestral spike, and the other for ancestral RBD, combined with sVNT. The model is run for 4,000 steps with 2,000 burn-in for 4 chains. The convergence diagnostics indicate good mixing and convergence, with effective sample sizes (ESS) ranging from 1,000 to 6,500 and Potential Scale Reduction Factor (PSRF) <1.1 for all parameters. The code for this analysis and for the figures of the manuscript and appendix can be found at https://github.com/dchodge/covidbcell.

## Data Availability

All data produced are available online at https://github.com/dchodge/covidbcell

## Acknowledgements

Thank you to A Jessica Hadiprodjo, Dilini Rathnayake, Honghua Ding, and Louise Randall for helping with study procedures, sample collection and processing.

## Author contributions

DH: Conceptualisation, data curation, formal analysis, methodology, software, visualisation, writing—original draft, writing—review and editing.

LY: Validation, investigation, resources.

LC: Validation, investigation, resources.

SM: Funding acquisition, writing—review and editing.

KS: Funding acquisition, writing—review and editing.

SGS: Conceptualisation, project administration, funding acquisition, writing—review and editing.

AF: Data Curation, Conceptualisation, project administration, funding acquisition, writing— review and editing.

AK: Conceptualisation, project administration, funding acquisition, writing—review and editing.

## Competing interest statement

DH: None

LY: None

LC: None

SM: None

KS: None

SGS: Reports advisory board participation and/or consulting for Moderna, Pfizer, Novavax,

CSL Seqirus and Sanofi.

AF: Funding from Sanofi AK: None

## Funding sources

This work was supported by the National Institutes of Health [R01AI141534 to SGS, AF, AJK, DH]. The WHO Collaborating Centre for Reference and Research on Influenza is funded by the Australian Government Department of Health. This work was also supported by the National Institute of Allergy and Infectious Diseases, National Institutes of Health, Department of Health and Human Services Centers of Excellence for Influenza Research and Response [CEIRR] grant number HHSN272201400005C to the University of Rochester and a subcontract to KS.

## Notes

### Author Declarations

The Royal Melbourne Hospital Human Research and Ethics Committee (HREC/63096/MH-2020) of Royal Melbourne Hospital gave ethical approval for this work. The Royal Melbourne Hospital Human Research Ethics Committee (HREC/54245/MH-2019) and LSHTM Observational Research Ethics Committee of London School of Hygiene and Tropical Medicine gave ethical approval for the use of this data for analysis.

## REFERENCES

1. T. Fiolet, Y. Kherabi, C.-J. MacDonald, J. Ghosn, N. Peiffer-Smadja, Comparing COVID-19 vaccines for their characteristics, efficacy and effectiveness against SARS-CoV-2 and variants of concern: a narrative review. Clin. Microbiol. Infect. 28, 202–221 (2022).

2. Polack Fernando P., et al., Safety and Efficacy of the BNT162b2 mRNA Covid-19 Vaccine. N. Engl. J. Med. 383, 2603–2615 (2020).

3. Thomas Stephen J., et al., Safety and Efficacy of the BNT162b2 mRNA Covid-19 Vaccine through 6 Months. N. Engl. J. Med. 385, 1761–1773 (2021).

4. M. Voysey, et al., Safety and efficacy of the ChAdOx1 nCoV-19 vaccine (AZD1222) against SARS-CoV-2: an interim analysis of four randomised controlled trials in Brazil, South Africa, and the UK. Lancet 397, 99–111 (2021).

5. J. Wei, et al., Comparative effectiveness of BNT162b2 and ChAdOx1 nCoV-19 vaccines against COVID-19. BMC Med. 21, 78 (2023).

6. K. Shioda, et al., Comparative effectiveness of alternative intervals between first and second doses of the mRNA COVID-19 vaccines. Nat. Commun. 15, 1214 (2024).

7. N. Ahmed, et al., To be remembered: B cell memory response against SARS-CoV-2 and its variants in vaccinated and unvaccinated individuals. Scand. J. Immunol. 99, e13345 (2024).

8. Y. Liu, et al., Superior immunogenicity of mRNA over adenoviral vectored COVID-19 vaccines reflects B cell dynamics independent of anti-vector immunity: Implications for future pandemic vaccines. Vaccine 41, 7192–7200 (2023).

9. K. J. Ewer, et al., T cell and antibody responses induced by a single dose of ChAdOx1 nCoV-19 (AZD1222) vaccine in a phase 1/2 clinical trial. Nat. Med. 27, 270–278 (2021).

10. M. Müller, et al., Single-dose SARS-CoV-2 vaccinations with either BNT162b2 or AZD1222 induce disparate Th1 responses and IgA production. BMC Med. 20, 29 (2022).

11. V. G. Hall, et al., Delayed-interval BNT162b2 mRNA COVID-19 vaccination enhances humoral immunity and induces robust T cell responses. Nat. Immunol. 23, 380–385 (2022).

12. N. D. Almeida, et al., The effect of dose-interval on antibody response to mRNA COVID-19 vaccines: a prospective cohort study. Front. Immunol. 15, 1330549 (2024).

13. I. Garcia-Fogeda, et al., Within-host modeling to measure dynamics of antibody responses after natural infection or vaccination: A systematic review. Vaccine 41, 3701–3709 (2023).

14. A. M. Smith, Decoding immune kinetics: unveiling secrets using custom-built mathematical models. Nat. Methods 21, 744–747 (2024).

15. I. Balelli, et al., A model for establishment, maintenance and reactivation of the immune response after vaccination against Ebola virus. J. Theor. Biol. 495, 110254 (2020).

16. F. X. Heinz, K. Stiasny, Distinguishing features of current COVID-19 vaccines: knowns and unknowns of antigen presentation and modes of action. NPJ Vaccines 6, 104 (2021).

17. S. P. Weisberg, et al., Distinct antibody responses to SARS-CoV-2 in children and adults across the COVID-19 clinical spectrum. Nat. Immunol. 22, 25–31 (2021).

18. A. C. Dowell, et al., Children develop robust and sustained cross-reactive spike-specific immune responses to SARS-CoV-2 infection. Nat. Immunol. 23, 40–49 (2022).

19. D. A. Collier, et al., Age-related immune response heterogeneity to SARS-CoV-2 vaccine BNT162b2. Nature 596, 417–422 (2021).

20. J. Wei, et al., Antibody responses to SARS-CoV-2 vaccines in 45,965 adults from the general population of the United Kingdom. Nat Microbiol 6, 1140–1149 (2021).

21. A. Nicolas, et al., An extended SARS-CoV-2 mRNA vaccine prime-boost interval enhances B cell immunity with limited impact on T cells. iScience 26, 105904 (2023).

22. Z. Wang, et al., Naturally enhanced neutralizing breadth against SARS-CoV-2 one year after infection. Nature 595, 426–431 (2021).

23. M. Sakharkar, et al., Prolonged evolution of the human B cell response to SARS-CoV-2 infection. Sci Immunol 6 (2021).

24. A. Sette, S. Crotty, Immunological memory to SARS-CoV-2 infection and COVID-19 vaccines. Immunol. Rev. 310, 27–46 (2022).

25. P. Dogra, et al., A modeling-based approach to optimize COVID-19 vaccine dosing schedules for improved protection. JCI Insight 8 (2023).

26. K. A. Pape, et al., High-affinity memory B cells induced by SARS-CoV-2 infection produce more plasmablasts and atypical memory B cells than those primed by mRNA vaccines. Cell Rep. 37, 109823 (2021).

27. J. S. Turner, et al., SARS-CoV-2 mRNA vaccines induce persistent human germinal centre responses. Nature 596, 109–113 (2021).

28. R. Keeton, et al., Impact of SARS-CoV-2 exposure history on the T cell and IgG response. Cell Rep Med 4, 100898 (2023).

29. J. S. Tsang, et al., Improving Vaccine-Induced Immunity: Can Baseline Predict Outcome? Trends Immunol. 41, 457–465 (2020).

30. M. R. Castrucci, Factors affecting immune responses to the influenza vaccine. Hum. Vaccin. Immunother. 14, 637–646 (2018).

31. D. Gouglas, M. Christodoulou, R. Hatchett, The 100 Days Mission—2022 Global Pandemic Preparedness Summit. Emerg. Infect. Dis. 29 (2023).

32. C. W. Tan, et al., A SARS-CoV-2 surrogate virus neutralization test based on antibody-mediated blockage of ACE2-spike protein-protein interaction. Nat. Biotechnol. 38, 1073–1078 (2020).

