## Supplementary Information for "Memory B cell proliferation drives differences in neutralising responses between ChAdOx1 and BNT162b2 SARS-CoV-2 vaccines"

Supplementary information to the article “Memory B cell proliferation drives differences in neutralising responses between ChAdOx1 and BNT162b2 vaccines”

Overview

This document presents a Bayesian model that describes antibody production in response to vaccination. The model captures the dynamic interplay between various immune markers, including memory B cells, plasma cells, plasmablasts, and antibodies. By employing ordinary differential equations (ODEs), we delineate the complex relationships among these immune components and explore how individual host factors contribute to the variability in immune responses.

The Bayesian framework is particularly advantageous in this context because it allows for the specification of intricate non-linear model structures. Moreover, it enables us to infer credible intervals for key parameters of interest, especially those that are crucial in understanding the in-host immune response. This approach provides a robust statistical foundation for evaluating the uncertainty and variability inherent in biological systems, thereby offering deeper insights into the mechanisms driving immune responses post-vaccination.

CONTENTS

1. OUTLINE OF THE DATA
   1.1. Calibration dataset
   1.2. Validation dataset
   1.3. Testing of serological samples
2. GOALS OF THE ANALYSIS
3. DESCRIPTION OF MODEL
   3.1. MATHEMATICAL MODEL OF HUMORAL IMMUNITY
   3.2. HIERARCHICAL EFFECTS
   3.3. LIKELIHOOD FUNCTION
   3.4. PRIORS
4. IMPLEMENTATION
   4.1. Software
   4.2. MCMC chain convergence and resolution for Ancestral spike
   4.3. MCMC chain convergence and resolution for Ancestral spike
5. POSTERIOR DISTRIBUTIONS
   5.1. Posterior predictive checks on the calibration dataset
   5.2. Posterior prediction on the validation dataset
   5.3. Posterior distributions of the fitted parameters.
   5.4. Drivers of memory B cell proliferation and antibody production
1. OUTLINE OF THE DATA

Our data consists of individual-level longitudinal serological samples taken before and following a second dose of SARS-CoV-2 vaccine. We consider two datasets, taken from two studies which ran concurrently and independently on distinct groups of individuals. Both studies were conducted in Australia during early 2021; a time when there was little SARS-CoV-2 transmission. As such, no individuals in either of the studies had confirmed clinical COVID-19 infections, and low transmission, paired with strict NPI measures, meant subclinical infections were unlikely to have occurred. Therefore, the population of both datasets is assumed to be infection-naive throughout the serological sampling.

1.1. *Calibration dataset*

A descriptive analysis of this dataset has been previously published.(Liu et al. 2023) It involves 41 individuals, of whom serological samples were taken before second dose, then at approximately days 7, 14, and 200 after vaccination. Due to the high frequency of samples taken close to vaccination date, it is possible to infer complex antibody kinetics immediately following vaccination. Therefore, this dataset is our *calibration dataset*.

1.2. *Validation dataset*

A descriptive analysis of this dataset is yet to be published. It involved 22 individuals, for whom serological samples were taken before the second dose, and then at day 30 post-vaccination. This dataset is used as a *validation dataset.* A summary of the host factors associated with the individuals I both studies is given in Table SM1.

1.3. *Testing of serological samples*

Although the serology between these two studies was conducted independently, the serological samples were tested according to the same protocol and were conducted in the same laboratory. Full details of the protocols can be found in the manuscript.

For each serological sample at each time point for each individual, we determined the concentration of memory B cells to RBD and spike (gating: CD19^+^IgD^-^AncetralRBD^+^CD71^+^/CD19^+^IgD^-^Ancetralspike^+^CD71^+^), the concentration plasmblasts to RBD and spike (gating: activated memory B-cell plus CD20^-^CD38^+^), and titre identified through sVNT titres. (**Figure SM1**). These B lymphocytes are measured as they are the main antibody-secreting cells (ASC) for a memory response, and thus combining these measures with the sVNT titres gives the data needed to establish a framework which relates antibody concentrations in serum and the B lymphocytes from which they are secreted. **Figure SM2** shows data showing the dynamics of these three biomarkers over time for all individuals, including RBD and spike for the calibration and validation dataset.

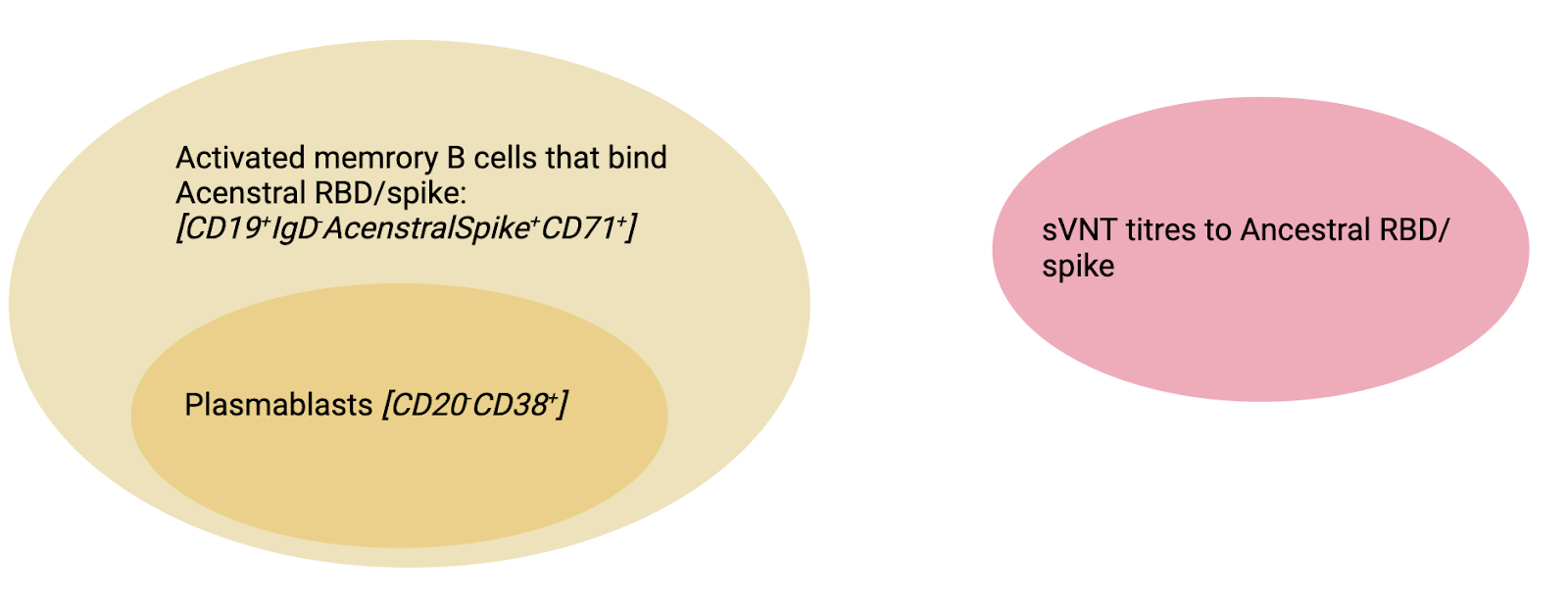

**Figure SM1.** Schematic showing the gating protocol for identifying MBC, PB and sVNT tires.

|  | Study A (Calibration) (N=41) | Study B (Validation) (N=22) | Overall (N=63) |
| --- | --- | --- | --- |
| **Vaccine type** |  |  |  |
| BNT162b2 | 33 (80.5%) | 11 (50.0%) | 44 (69.8%) |
| ChAdOx1 | 8 (19.5%) | 11 (50.0%) | 19 (30.2%) |
| **Time since first dose (days)** |  |  |  |
| <28 days | 26 (63.4%) | 9 (40.9%) | 35 (55.6%) |
| 28+ days | 15 (36.6%) | 13 (59.1%) | 28 (44.4%) |
| **Age (yrs)** |  |  |  |
| <30 years | 3 (7.3%) | 4 (18.2%) | 7 (11.1%) |
| 30–39 years | 15 (36.6%) | 6 (27.3%) | 21 (33.3%) |
| 40–49 years | 8 (19.5%) | 3 (13.6%) | 11 (17.5%) |
| 50–59 years | 13 (31.7%) | 6 (27.3%) | 19 (30.2%) |
| 60+ years | 2 (4.9%) | 3 (13.6%) | 5 (7.9%) |

Table SM1. Summary of the host factors across both datasets.

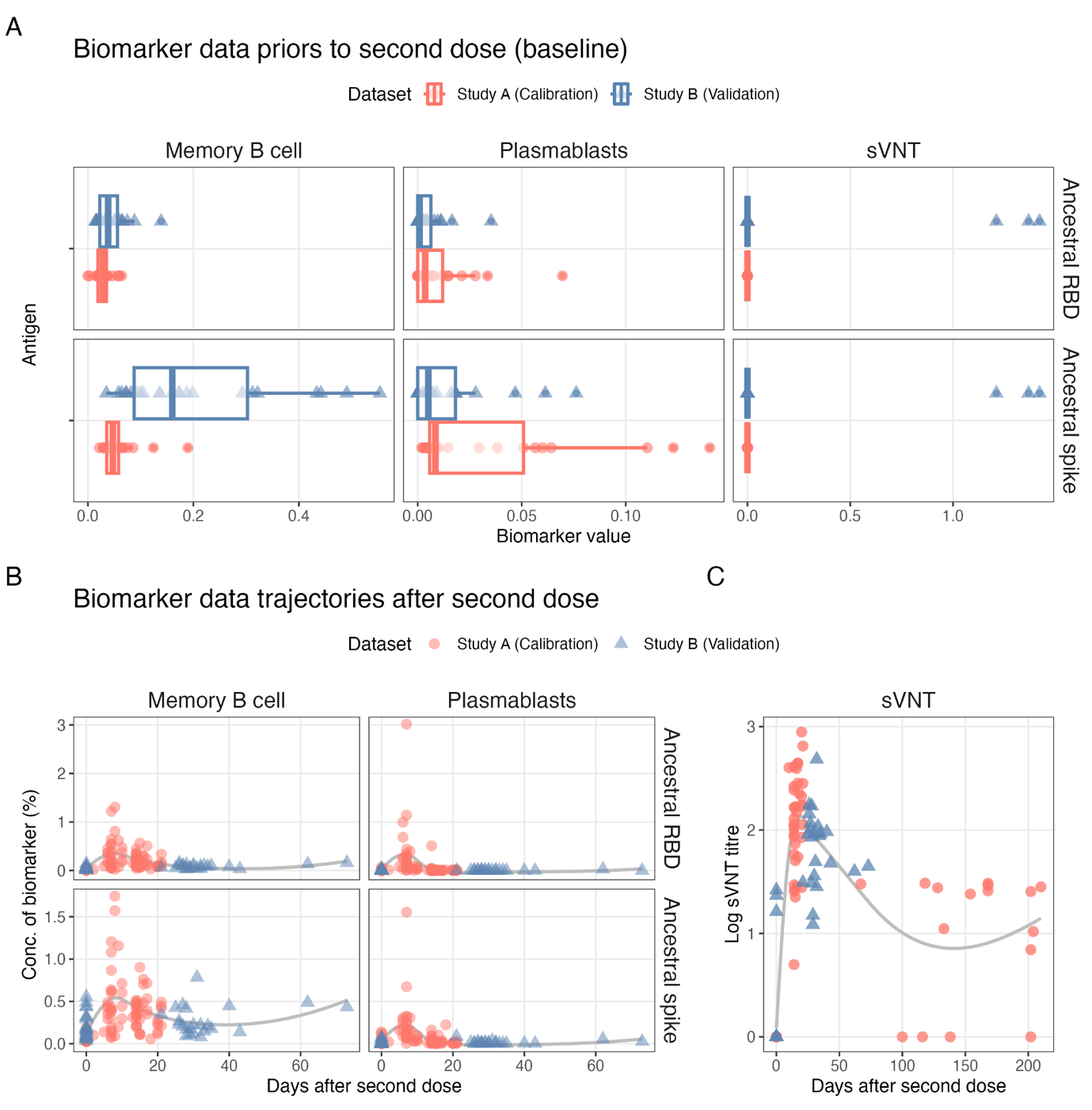

**Figure SM2.** Summary of the measured immune biomarker values at A) the baseline (prior to vaccination) and B) in response to vaccination for both the calibration dataset (red) and the validation dataset (blue).

2. GOALS OF THE ANALYSIS

We aim to develop a mechanistic model of antibody production to gain a deeper understanding of the relationship between antibody-secreting cells (ASC), such as plasma cells and plasmablasts, and the changes in serum antibody concentrations over time following a second dose of SARS-CoV-2 vaccination. Our objectives are threefold:

A. *Establish a mechanistic model of antibody production in response to COVID-19 vaccination*

We intend to create a dynamic system described by ordinary differential equations (ODEs) to illustrate the relationships between various immune components, memory B cells, plasmablasts, and antibody concentration (measured by sVNT titres). This mechanistic model aims to enhance our understanding of the immune response dynamics post-vaccination. By predicting immune responses based on individual-level baseline immune measures, we will evaluate the model's predictive accuracy using the Continuous Ranked Probability Score (CRPS) against an unseen validation dataset. Insights gained from this model can guide the design of vaccines that elicit stronger and longer-lasting antibody responses, thereby improving vaccine efficacy.

B. Determine the host factors driving immune heterogeneity to vaccination

Incorporating hierarchical effects into the model will help us assess the impact of host factors on the kinetics of memory B cells and antibodies. Identifying how previous vaccination history—considering vaccine type and timing—affects immune responses will inform improvements in vaccine formulations. Understanding the variability in immune responses due to host factors will enable personalised vaccination strategies to enhance overall vaccine effectiveness.

C. *Determine the temporal variation in the origin of antibody production over time*

By analysing the kinetics within the fitted dynamic models, we aim to understand the timeline of antibody production, including the transition from short-term to long-term immunity and the roles of different immune cells and organs over time. This temporal analysis will help predict the duration of immunity provided by the vaccine, thereby informing public health policies on optimal vaccination schedules and the frequency of booster doses.

3. DESCRIPTION OF MODEL
   3.1 MATHEMATICAL MODEL OF HUMORAL IMMUNITY

For the measured biomarkers, we devised an in-host mathematical model of humoral immunity to assess the kinetics of antibody production for each vaccine type. Figure SM3 schematically shows the underlying biological process relating to vaccine antigen, B-cell proliferation, differentiation, and antibody secretion from plasmablasts and plasma cells**.**

We assume that the vaccine antigen stimulates the proliferation of memory B cells. After this stimulation, these cells differentiate along one of two pathways. They can either become short-lived plasmablasts, which secrete an initial burst of antibodies in response to vaccination, or they can migrate to the germinal center. In the germinal center, they remain for approximately two weeks before differentiating into long-lived plasma cells, which also secrete antibodies.

In our serological samples, we measure the concentration of memory B cells and plasmablasts that respond to the ancestral antigen (spike or RBD). Additionally, we measure the concentration of antibodies against the ancestral variant using surrogate virus neutralization tests (sVNT).

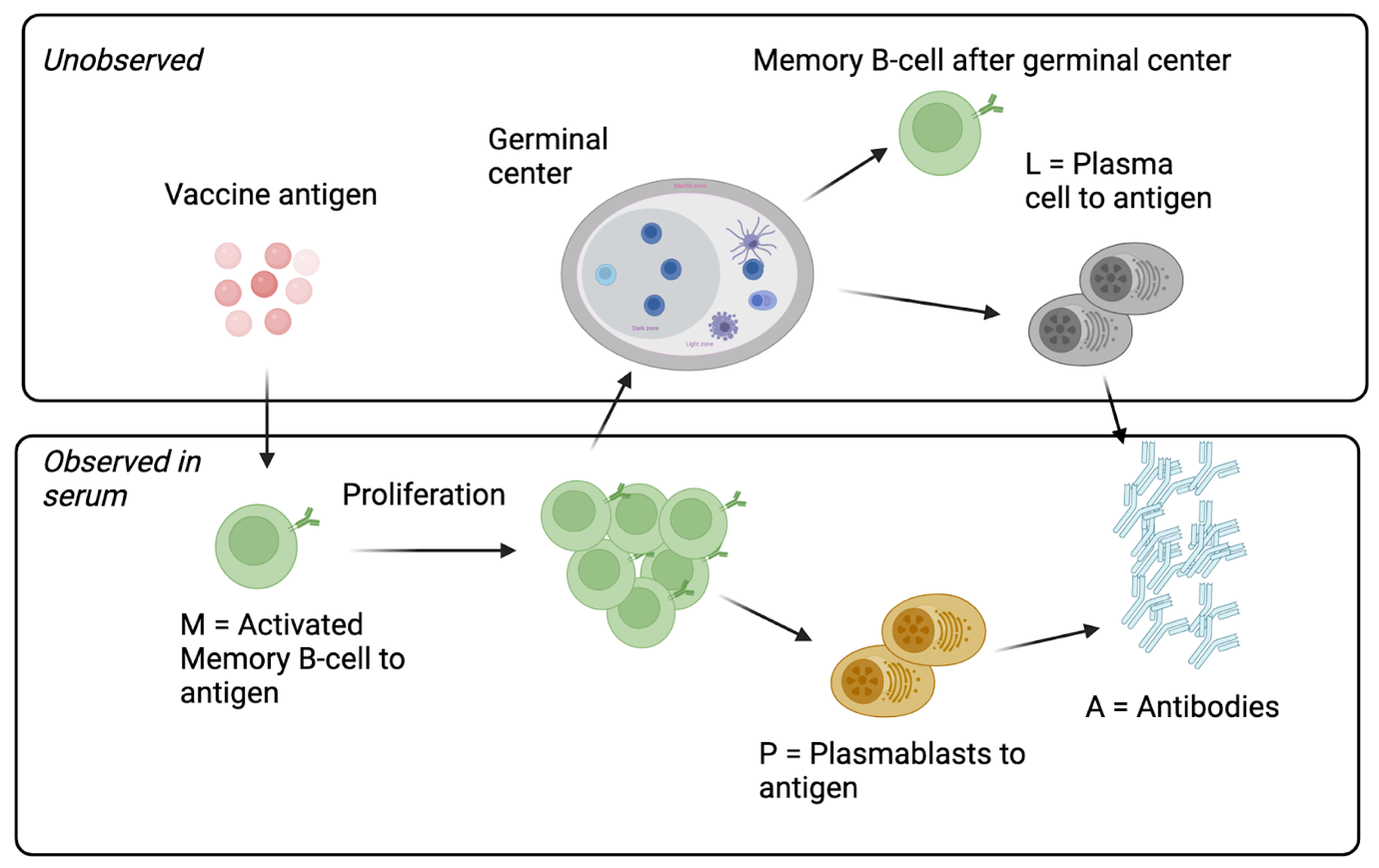

**Figure SM3.** Schematic of the underlying immunological mechanism leading to the production of antibodies to SARS-CoV-2.

Converting this immunological pathway into a dynamic system, we construct a compartmental model given in Figure SM4, noting the nonlinear nature of the assumed relationships between these biomarkers. The resulting differential equations associated with this dynamic model are also given in Figure SM4. The state variables and parameters associated with this system of ODEs are given in Tables SM2 and SM3.

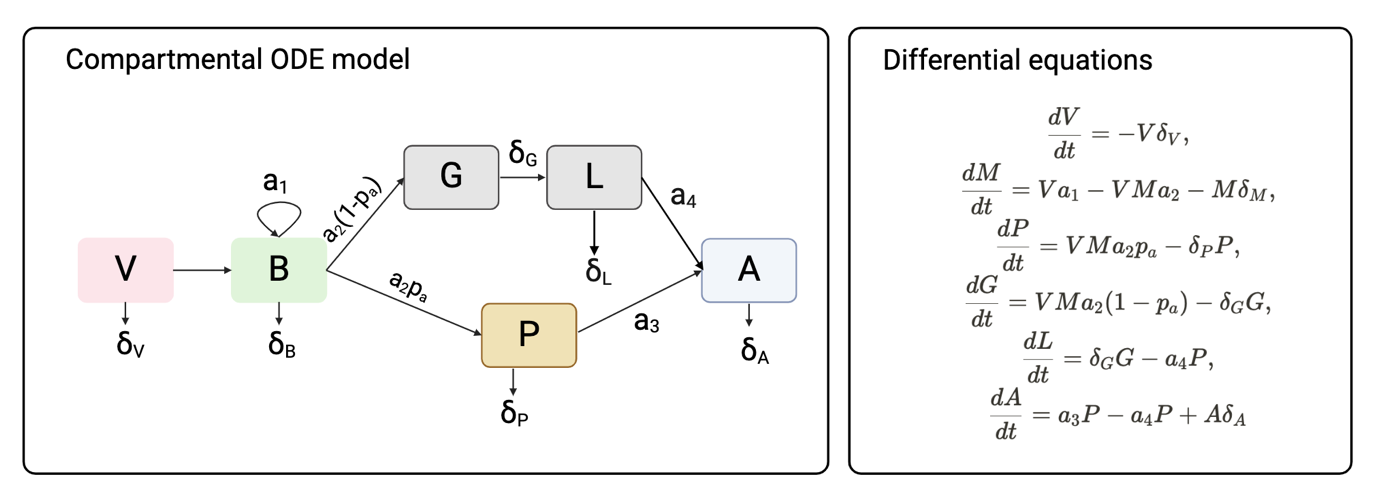

**Figure SM4.** Schematic of the underlying immunological mechanism leading to the production of antibodies to SARS-CoV-2.

| State variable symbol | Description | Data |
| --- | --- | --- |
| V | Concentration of SARS-CoV-2 antigen in vaccine | N/A* |
| B | Proportion of memory B cell which are activated and bind to Ancestral RBD/spike (and not plasmablasts) | See Figure SM1 |
| P | Proportion of memory B cell which are plasmablasts and bind to Ancestral RBD/spike | See Figure SM1 |
| G | Proportion of activated B cells in germinal center | *Latent* |
| L | Amount of plasma cells created from germinal center | *Latent* |
| A | Concentration of antibodies which neutralise ancestral RBD/spike | sVNT titres |

Table SM2. State variables. *Integrates out in Equations X.

| Parameter symbol | Description | Dimension |
| --- | --- | --- |
| $\delta_{V}$ | Rate of decay of vaccine antigen | day^-1^ |
| $\delta_{B}$ | Rate of decay of memory B-cells | day^-1^ |
| $\delta_{P}$ | Rate of decay of plasmablasts | day^-1^ |
| $\delta_{G}$ | Time for germinal centres to produce affinity maturated plasma cells | day^-1^ |
| $\delta_{L}$ | Rate of decay of plasma cells | day^-1^ |
| $\delta_{A}$ | Rate of decay of neutralizing antibodies | day^-1^ |
| a_1_ | **Immunogenicity of vaccine**  Rate of proliferation of memory B-cells per vaccine dose per day | day^-1^ |
| a_2_ | Rate of differentiation of memory B-cells to plasmablasts/plasma cells per vaccine dose per day | day^-1^ |
| a_3_ | **Affinity of vaccine-induced antibodies from plasmablasts**  Rate of production of neutralizing antibodies per conc. of plasmablasts per day | log_2_(ab)day^-1^ |
| a_4_ | **Affinity of vaccine-induced antibodies from plasma cells**  Rate of production of neutralizing antibodies per conc. of plasma cells per day | log_2_(ab)day^-1^ |
| p_1_ | Proportion of differentiated memory B-cells which become plasmablasts | *None* |

Table SM3. Summary of parameters in the ODE model

3.2 HIERARCHICAL EFFECTS

We want to assess the influence of vaccine type and host factors (age and time since vaccination) on the kinetics of antibody production. To do this, we add hierarchical effects to our parameters of interest: the rate of proliferation of memory B-cells per vaccine dose per day (a_1_), rate of production of neutralising antibodies per conc. of plasmablasts per day, (a_3_) and the rate ate of production of neutralising antibodies per conc. of plasma cells per day (a_4_). The covariates we consider in our hierarchical effects are the vaccine type (CdAdOx1, BNT162b2), age groups (<30, 30–39, 40–49, 50–59, 60+), time since last vaccination (<28 days, 28+days). Finally, we incorporate individual-level effects to each parameter to account for the high amount of individual-level variability between each individual response, reducing biases from unusual kinetics and improving the model's fit. We incorporate these into the mathematical model through the equations.

$$a_{1}= \beta_{1}+\nu_{1,j}x_{v,i}+\alpha_{1,j}x_{a,i}+\tau_{1,l}x_{t,i}+\rho_{1,l}x_{i},$$

$$a_{3}= \beta_{3}+\nu_{3,j}x_{v,i}+\alpha_{3,j}x_{a,i}+\tau_{3,l}x_{t,i}+\rho_{3,l}x_{i},$$

$$a_{4}= \beta_{4}+\nu_{4,j}x_{v,i}+\alpha_{4,j}x_{a,i}+\tau_{4,l}x_{t,i}+\rho_{4,l}x_{i}$$

**Equations 1. Hierarchical equations**

The definition of the hierarchical parameters is given in **Table SM4.**

3.3. LIKELIHOOD FUNCTION

We assume that the measured biomarkers are subject to an error which follows a normal distribution. Therefore, the biomarker-dependent likelihood, which relates the biomarker data (memory B cell (B), plasmablasts (P) and antibodies (A)) and the model-predicted biomarker quantity is given by:

$$p(Y_{B}|X_{B}, \sigma_{B}) = \prod_{i = 1}^{N} \prod_{t_{i}=1}^{T_{i}} \frac{1}{\sigma_{B}\sqrt{2\pi}}exp\left( -\frac{(Y_{B, t_{i}}-{X_{B}(t_{i}))}^{2}}{2\sigma_{B}^{2}} \right)$$

$$p\left( Y_{P} | X_{P},\sigma_{P} \right)=\prod_{i = 1}^{N} \prod_{t_{i}=1}^{T_{i}} \frac{1}{\sigma_{P}\sqrt{2\pi}}exp\left( -\frac{(Y_{P, t_{i}}-{X_{P}(t_{i}))}^{2}}{2\sigma_{P}^{2}} \right)$$

$$p\left( Y_{A} | X_{A},\sigma_{A} \right)=\prod_{i = 1}^{N} \prod_{t_{i}=1}^{T_{i}} \frac{1}{\sigma_{A}\sqrt{2\pi}}exp\left( -\frac{(Y_{A, t_{i}}-{X_{A}(t_{i}))}^{2}}{2\sigma_{A}^{2}} \right)$$

With a combined likelihood given by

$$L(Y | X) = p\left( Y_{B} | X_{B},\sigma_{B} \right)p\left( Y_{P} | X_{P},\sigma_{P} \right) p\left( Y_{A} | X_{A},\sigma_{A} \right)$$

**Equations 2.** Likelihood equations.

Where$X_{P}(t_{i})$, $X_{B}(t_{i})$, $X_{A}(t_{i})$, is the model-predicted value of the memory B cells, plasmablasts, and antibodies at time t_i_ and $Y_{B, t_{i}}$, $Y_{P, t_{i}}$and $Y_{A, t_{i}}$ is the corresponding data at that time point. The values $\sigma_{B},$ $\sigma_{P}$ and $\sigma_{A}$ are fitted values for the standard deviations for each immune marker.

The model-predicted values for memory B cells, plasmablasts, and antibodies are estimates from the ODE system outlined in Figure SM. That is by integrating;

$$\frac{dX}{dt}=f(X, \theta, t)$$

Where X(t) = {$X_{B}, X_{P}, X_{G}, X_{L}, X_{A}$} are the state variables, and theta are the fitted parameter values and evaluating at the same time points as the data. *Note:* that we integrate out the equation for V and redefine the parameters as appropriate.

3.4 PRIORS

We identify three groups of parameters for which we need to describe prior distributions:

i) Parameters describing the dynamic system

For the $\delta_{k}$ values (k = {V, B, P, G, L, A}), which describe the average rates of degradation for the biomarkers in the model, we use strongly informed priors from immunological observations to derive a prior distribution for each. For the average rate of decay of vaccine antigen ($\delta_{V}$) we choose a uniform distribution between 1 and 30 days, assuming that detectable antigen is rarely seen in infected humans after 30 days.(Russell et al. 2023) As we are modelling vaccination and not infection, we use a non-informative uniform prior to account for translational issues between active viral degradation and vaccine antigen degradation. For the rate of degradation of memory B cells (MBC), we fix the value to 1000 days, reflecting the assumption that MBC is unlikely to degrade over the relatively short time span of the study (~30–200 days). For the average rate of degradation of plasmablasst we chose a strongly informed prior of *N*(4, 1) reflecting the belief that plasmablasts last around 3–5 days.(Khodadadi et al. 2019) For the average time spent in the Germinal Center, we use a prior of N18, 2) reflecting the belief that individuals spend between 14 –22 days in the germinal center before the creation of plasma cells.(Flowers and Eisenbarth 2024) The average rate of degradation of plasma cells is set at Normal(700, 200) reflecting the belief that they remain for over 1 year.(Khodadadi et al. 2019) The rate of decay of antibodies is N(30, 5), reflecting the belief that antibody persistent for an average one month (Foss et al. 2024).

Next, we describe the chosen priors for the parameters that define the MBC and antibody kinetics, a_1_, a_2_, a_3_, a_4_ and *p*_1._ For a_2_, rate of differentiation of memory B-cells to plasmablasts/plasma cells per vaccine dose per day, we choose a non-informative prior of U(0, 1), suggesting anywhere between 0 to 100% of the MBC differentiate per day. For a_1_, a_3_, and a_4_ we require positive values, but it is unclear what the upper bound of their distribution should be. By inspection of the prior prediction distribution, we chose prior distributions of a_1_ ~ U(0, 2), a_3_ ~ U(0, 6), a_4_ ~ U(0, 0.5), ensuring that the sVNT titres remain in feasible ranges.

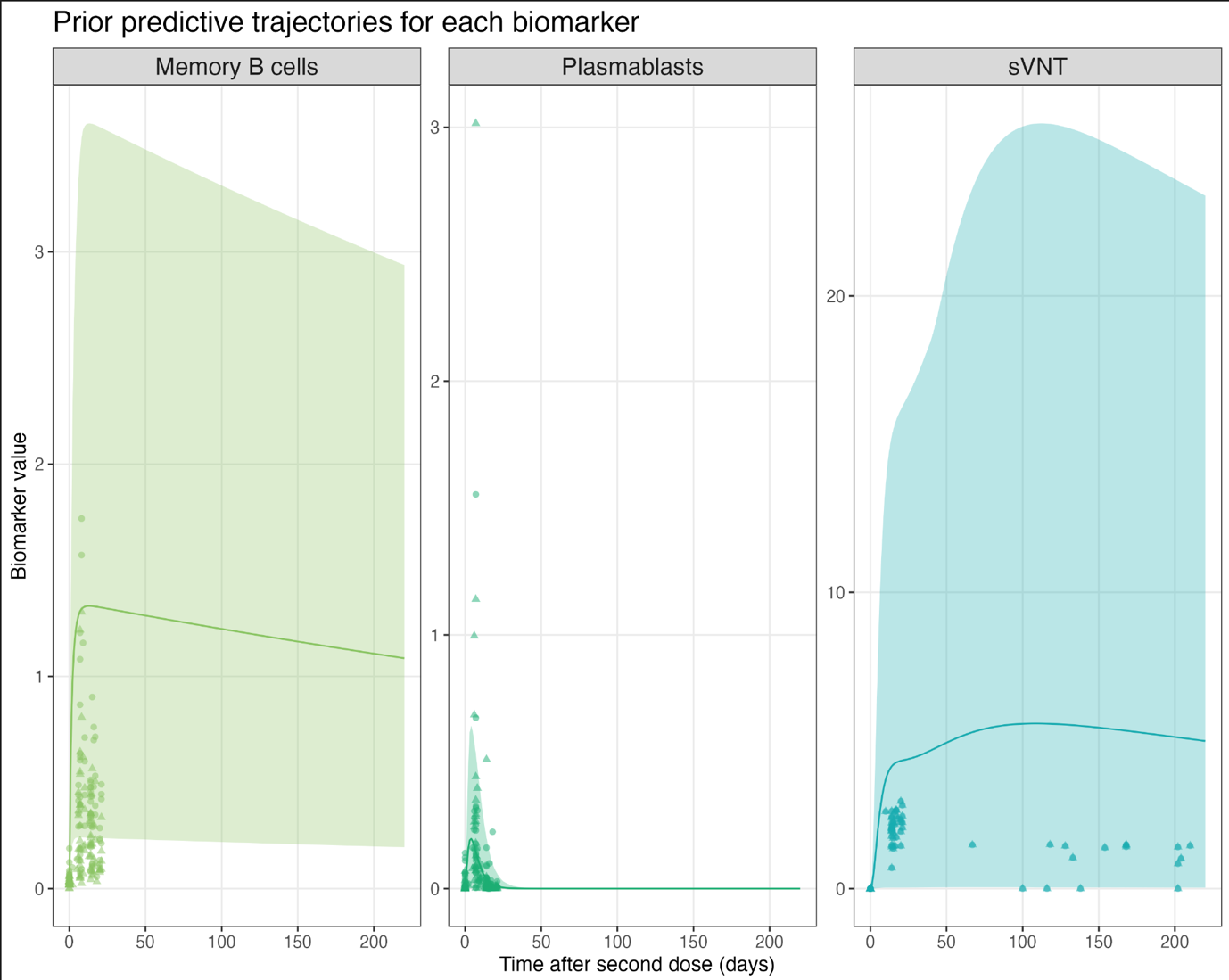

**Figure SM5.** Prior predictive distribution for the chosen priors in the model (line and area). The data for the spike and RBD for the validated is plotted through markers.

ii) Parameters of the hierarchical effects

In the implementation, we do not sample a_1_, a_3_, and a_4_ and instead determine them through **Equations 1.** We, therefore, must ensure that the distribution of a_1_, a_3_, and a_4_ reflect their chosen priors by carefully selecting the prior distributions of the hierarchical effects.

Recalling $\boldsymbol{Equations 1}$**.** We ensure the limits of the uniform distributions are realised by adding an inverse logit transformation:

$$a_{1}= g\left( \theta_{1} \right)= {logit}^{-1}(\beta_{1}+\nu_{1,j}x_{v,i}+\alpha_{1,j}x_{a,i}+\tau_{1,l}x_{t,i}+\rho_{1,l}x_{i})*b_{1}$$

$$a_{3}= g\left( \theta_{3} \right)= {logit}^{-1}(\beta_{3}+\nu_{3,j}x_{v,i}+\alpha_{3,j}x_{a,i}+\tau_{3,l}x_{t,i}+\rho_{3,l}x_{i})*b_{3}$$

$$a_{4}= g\left( \theta_{4} \right)= {logit}^{-1}(\beta_{4}+\nu_{4,j}x_{v,i}+\alpha_{4,j}x_{a,i}+\tau_{4,l}x_{t,i}+\rho_{4,l}x_{i})*b_{4}$$

**Equations 3. Inverse logit-transformed hierarchical effects**

Where b is the upper bound for a_1_, in this case b_1_ = 2, b_3_ = 6 and b_4_ = 0.5. Respecting the inverse logit transformation, we notice that if

$$q\sim N\left( 0,\sqrt{\frac{\pi}{3}} \right), p \sim{logit}^{-1}\left( 0,1 \right),$$

Then, the composition of q and p is approximately uniform, e.g.

$$p(q(x)) \sim U(0, 1)$$

As *q* is approximately equal to the logistic distribution with mean 0 and scale parameter 1. Therefore, by choosing a prior for $\beta_{1}$, $\beta_{3},$ and $\beta_{4}$~ $N\left( 0,\sqrt{\frac{\pi}{3}} \right)$, we can approximate a uniform distribution after the inverse logistic transformation.

The remaining parameters describing the effects of age, time since first dose, and vaccine type for a_1_, a_3_ and a_4_ are all implemented with non-centred parameterisation where pooling is given by $\eta\sigma$ where $\eta\sim N(0,1)$ and $\sigma\sim exponential(c)$. The value of c, which quantifies the partial poling, is determined by inspecting the inverse transformed values described by **Equations 3** to ensure they reflect that a_1_, a_3_, and a_4_ remain uniform after the addition covariates are added to the logistic transformation. **Figure S6** shows the results, and we chose p = 3 given the similarity between model A and B.

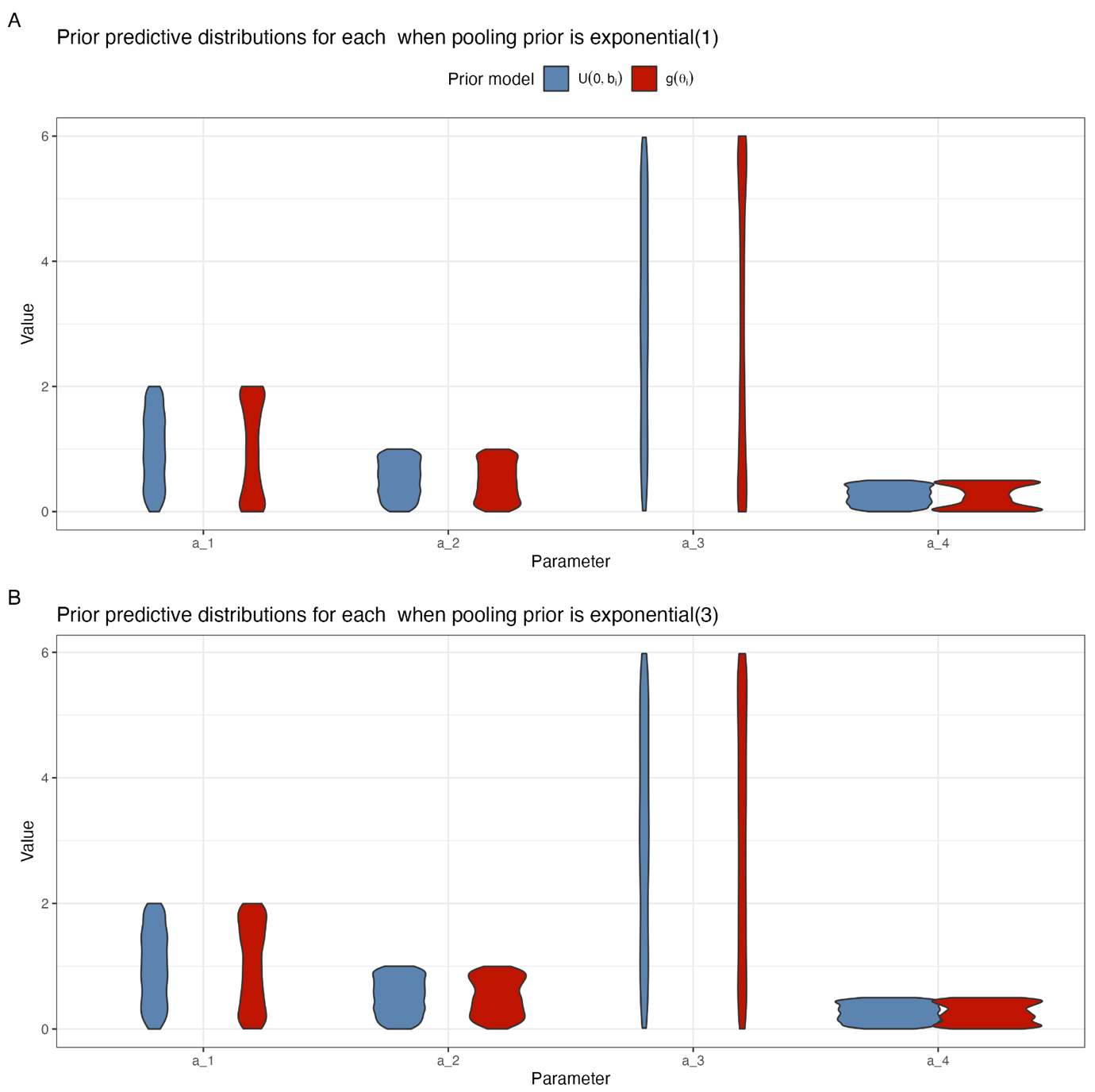

**Figure SM6. Comparison of the realised priors when i) a_i is sampled directly from a uniform (blue) and when ii) values of a_i are determined by sampling form the priors for the hierarchical effects (red).**

iii) Parameters describing the likelihood

There are three parameters: $\sigma_{B},$ $\sigma_{P}$ and $\sigma_{A}$ which describe the standard deviation of the normally distributed observation error for memory B cells, plasmablasts and antibody respectively. We use a weakly informative prior of exponential(1) for each parameter.

The priors for all the parameters can be found in Table SM4.

| **Symbol** | **Description** | **Prior** |
| --- | --- | --- |
| *Data* | | |
| $Y_{B, t_{i}}$ | Conc. of MBC at time t_i_  for individual *i*. | — |
| $Y_{A, t_{i}}$ | Conc. of PB at time t_i_  for individual *i*. | — |
| $Y_{A, t_{i}}$ | sVNT. of ancestral antibodies at time t_i_  for individual *i*. | — |
| x_v,i_ | Vaccine type for individual *i* | — |
| xa_,i_ | Age group for individual *i* | — |
| xt_,i_ | Dosing schedule group for individual *i* |  |
| *Dynamics ODE model of antibody production* | | |
| *a_1_* | Mean peak antibody boosting in study year *y* (log_2_) | Eq1.1 |
| a_2_ | Scale parameter in Gaussian process prior model for boosting | *U(0, 1)* |
| a_3_ | Length parameter in Gaussian process prior model for boosting | Eq1.1 |
| a_4_ | Intercepts for categorical age groups data, pooled effects | Eq1.1 |
| $\delta_{V}$ | Rate of decay of vaccine antigen | *U(1, 30)* |
| $\delta_{B}$ | Rate of decay of memory B-cells | *1000 days (fixed)* |
| $\delta_{P}$ | Rate of decay of plasmablasts | *N(4, 1)* |
| $\delta_{G}$ | Time for germinal centres to produce affinity maturated plasma cells | *N(18, 2)* |
| $\delta_{L}$ | Rate of decay of plasma cells | *N(730, 200)* |
| $\delta_{A}$ | Rate of decay of neutralizing antibodies | *N(30, 5)* |
| *p_1_* | Interaction term for categorical vaccine history data, pooled effects | *B(60, 40)* |
| *Hierarchical effects on a_1_* | | |
| $\beta_{1}$ | Intercept of *a_1_* | N$\left( 0,\sqrt{\frac{\pi}{3}} \right)$ |
| $\nu_{1,j}$ | Hierarchical effect of vaccine type *(j) o*n *a_1_* | N(0, sigma), sigma ~ *Exponential*(3) |
| $\alpha_{1,k}$ | Hierarchical effect of of age group *(k) o*n  *a_1_* | N(0, sigma), sigma ~ *Exponential*(3) |
| $\tau_{1,l}$ | Hierarchical effect of vaccine dosing schedule group (*l) o*n *a_1_* | N(0, sigma), sigma ~ *Exponential*(3) |
| $\rho_{1,j}$ | Inidividual-level hierarchical effect (i) on *a_1_* | N(0, sigma), sigma ~ *Exponential*(3) |
| *Hierarchical effects on a_3_* | | |
| $\beta_{3}$ | Intercept of *a_3_* | N$\left( 0,\sqrt{\frac{\pi}{3}} \right)$ |
| $\nu_{3,j}$ | Hierarchical effect of vaccine type *(j) o*n *a_3_* | N(0, sigma), sigma ~ *Exponential*(3) |
| $\alpha_{3,k}$ | Hierarchical effect of of age group *(k) o*n *a_3_* | N(0, sigma), sigma ~ *Exponential*(3) |
| $\tau_{3,l}$ | Hierarchical effect of vaccine dosing schedule group (*l) o*n *a_3_* | N(0, sigma), sigma ~ *Exponential*(3) |
| $\rho_{3,j}$ | Individual-level hierarchical effect (i) on *a_3_* | N(0, sigma), sigma ~ *Exponential*(3) |
| *Hierarchical effects on a_4_* | | |
| $\beta_{4}$ | Intercept of *a_4_* | N$\left( 0,\sqrt{\frac{\pi}{3}} \right)$ |
| $\nu_{4,j}$ | Hierarchical effect of vaccine type *(j) o*n *a_4_* | N(0, sigma), sigma ~ *Exponential*(3) |
| $\alpha_{4,k}$ | Hierarchical effect of of age group *(k) o*n *a_4_* | N(0, sigma), sigma ~ *Exponential*(3) |
| $\tau_{4,l}$ | Hierarchical effect of vaccine dosing schedule group (*l) o*n *a_4_* | N(0, sigma), sigma ~ *Exponential*(3) |
| $\rho_{4,j}$ | Individual-level hierarchical effect (i) on *a_4_* | N(0, sigma), sigma ~ *Exponential*(3) |
| *Likelihood function* | | |
| $\sigma_{B}$ | Standard deviation of normal distribution of observation model for MBC | *Exponential(1)* |
| $\sigma_{P}$ | Standard deviation of normal distribution of observation model for PB | *Exponential(1)* |
| $\sigma_{A}$ | Standard deviation of normal distribution of observation model for sVNT titre | *Exponential(1)* |

**Table SM4.** Summary of all the parameters and prior distributions in the Bayesia n model.

4. IMPLEMENTATION

*Note the naming convention for Figures changes for this point as they are referenced in the manuscript.*

4.1 Software

The model is coded and fitted using HMC via stan using cmdnstanr version 0.5.3 in R version 4.2.3. We solved the ODEs at each Markov chain step using the built in runge-kutta 4 method ‘ode_rk45’. A script for the stan code is given at <https://github.com/dchodge/covidbcell>.

The HMC was ran on two different datasets, one using the MBC and plasmablasts to Ancestral spike combined with the sVNT titres, and the other the MBC and plasmablasts to Ancestral RBD combined with the sVNT titres. We report the convergence and the resolution of these model fits separately.

4.2 MCMC chain convergence and resolution for Ancestral spike

The model is run for 4,000 steps, with 2,000 burn-in for four chains. The number of divergent transitions is 2/4000 (<0.1%), none of the chains hit the maximum tree depth of 10. The chains converged, with all parameters seeing a Potential Scale Reduction Factor (PSRF) of < 1.1, and mixed well; with Effective Sample Size (ESS) of tail and bulk ranging between 1,000 and 6,500 for all parameters. (See Figures below)

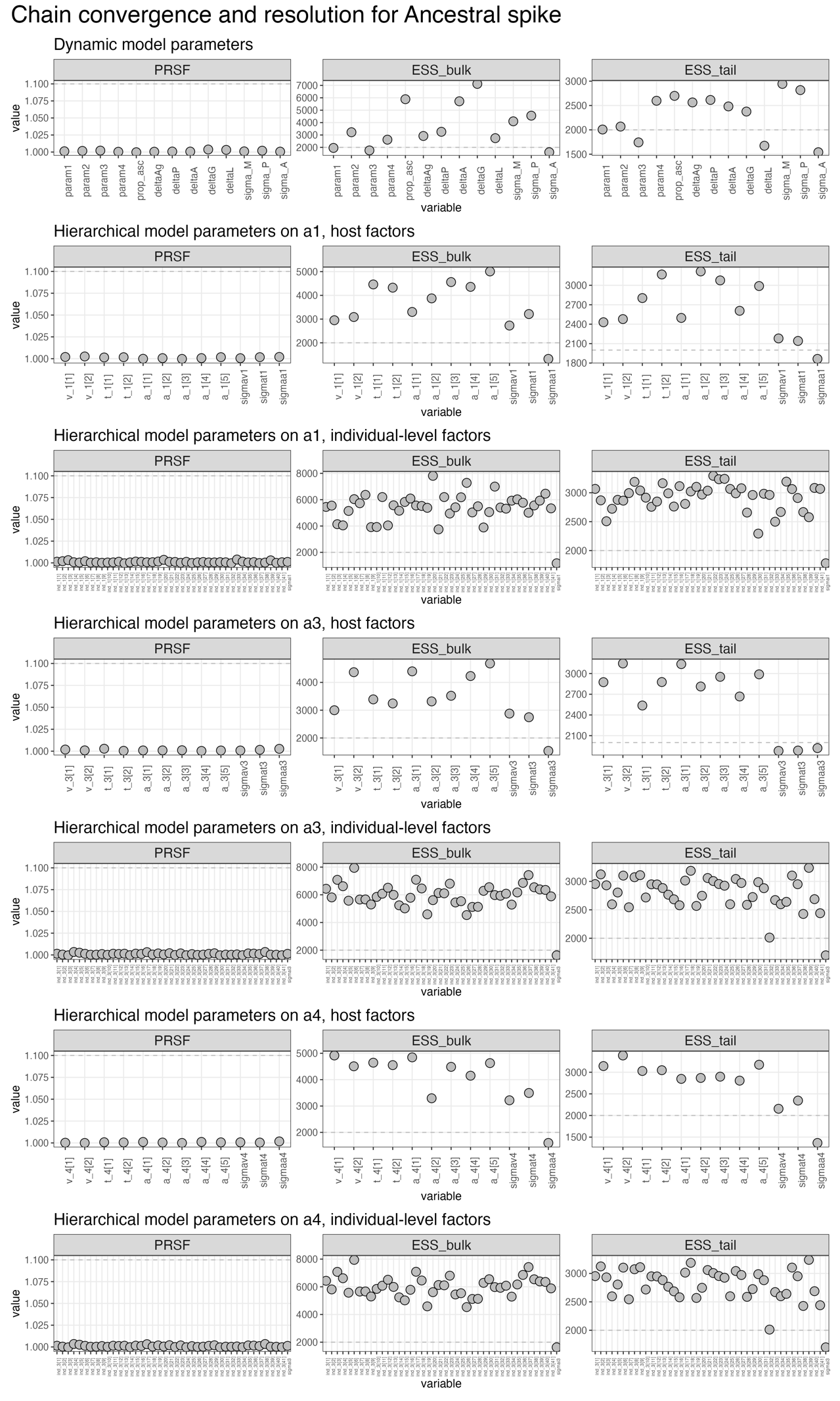

**Figure S1.** Plots showing the PRSF and ESS bulk and tail for all parameters fitted in the HMC sampler using data for Ancestral spike.

4.3 MCMC chain convergence and resolution for Ancestral spike

The model is ran for 4,000 steps, with 2,000 burn-in for 4 chains. The number of divergent transitions is 12/4000 (0.3%), none of the chains hit the maximum tree depth of 10. The chains converged, with all parameters seeing a Potential Scale Reduction Factor (PSRF) of < 1.1, and mixed well; with Effective Sample Size (ESS) of tail and bulk ranging between 1,000 and 6,500 for all parameters.

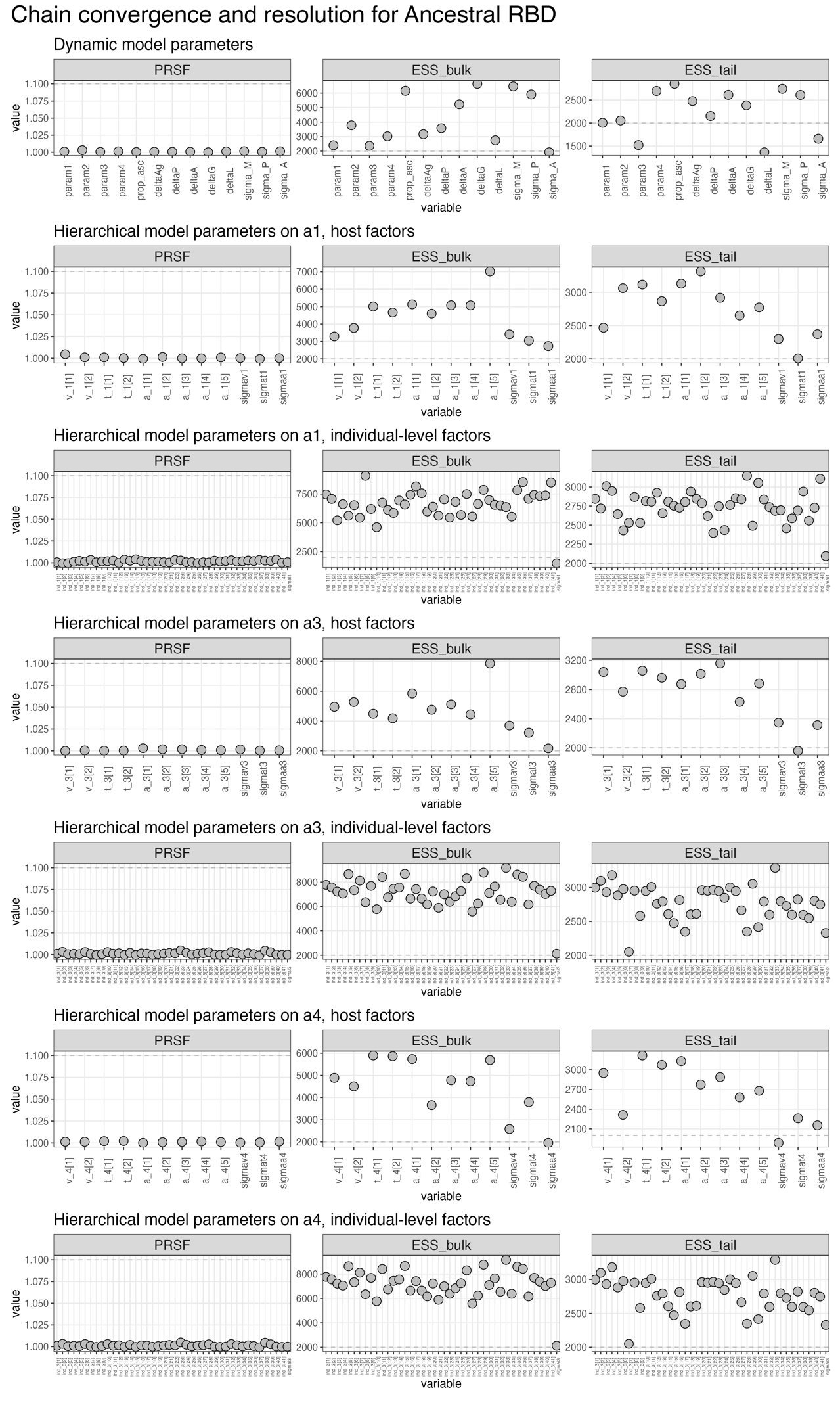

**Figure S2.** Plots showing the PRSF and ESS bulk and tail for all parameters fitted in the HMC sampler using data for Ancestral RBD.

5. POSTERIOR DISTRIBUTIONS

In reporting prior distributions, we report the central tendency and credible interval/posterior predictive interval (CrI/PPI) based on the median and the 95% equal-tailed distribution (ETI) i.e. ensuring 2.5% of the distribution is excluded at both ends.

5.1. Posterior predictive checks on the calibration dataset

To assess the suitability of the model fit, we plot the measure biomarker data, and $Y_{B, t_{i}}$, $Y_{P, t_{i}}$and $Y_{A, t_{i}}$ against the posterior predictive distributions of $X_{P}(t_{i})$, $X_{B}(t_{i})$, $X_{A}(t_{i}).$

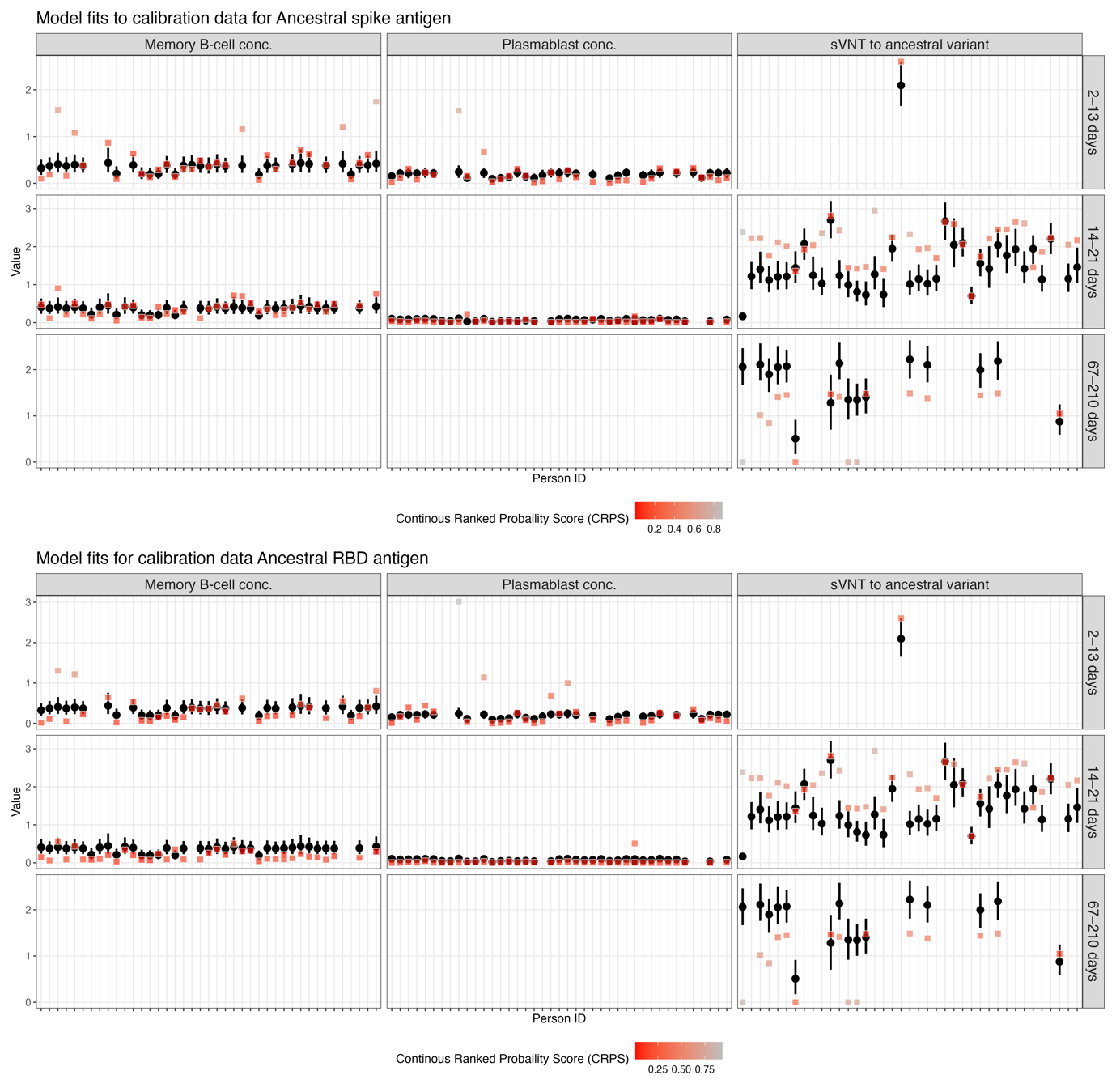

**Figure S3.** Comparison of the data and the model-predicted values for ancestral spike.

The individual-level fits for spike and RBD model on the calibration data for all three biomarkers are shown in Figure x.

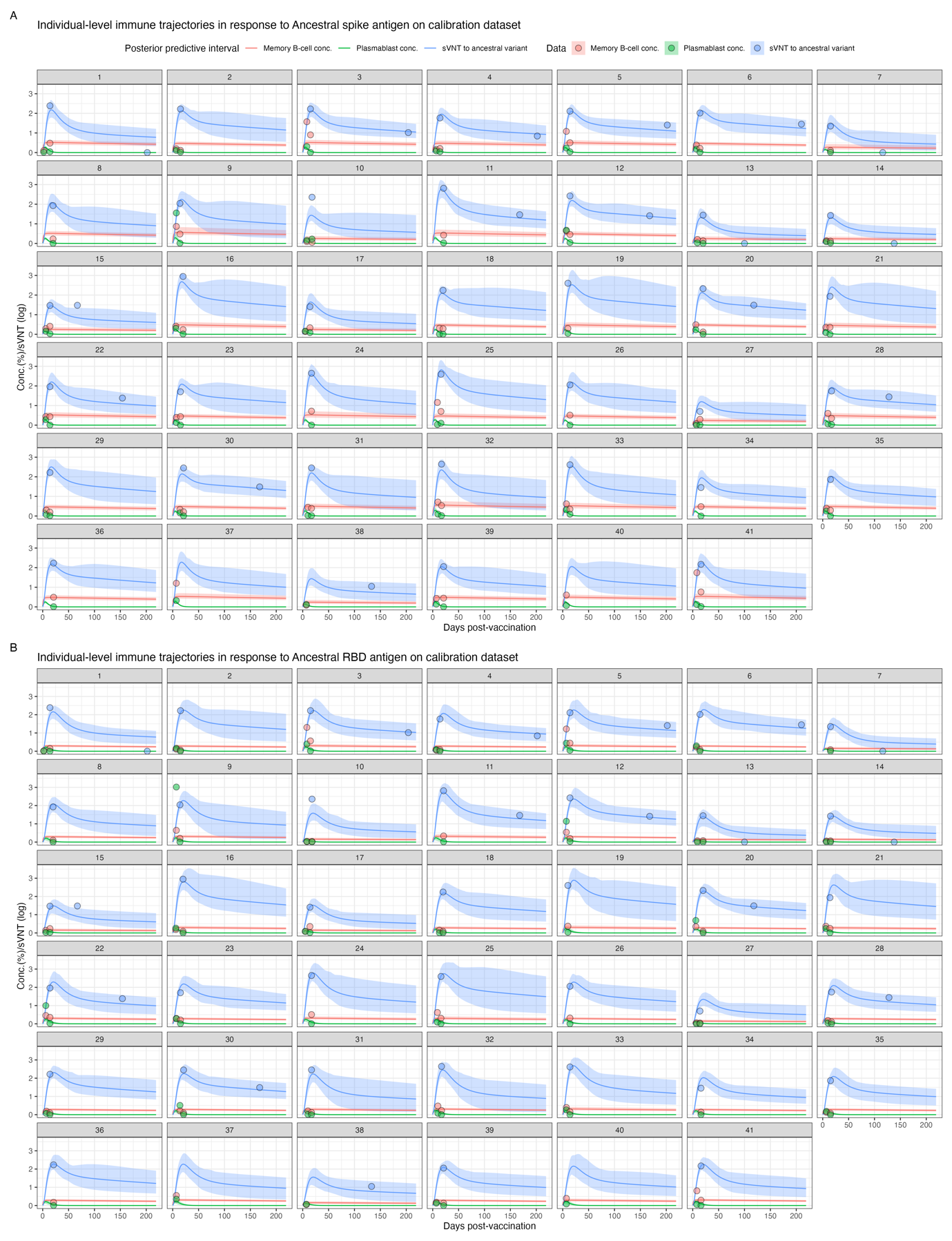

**Figure S4**. Individual-level trajectories comparing the data and the model-predicted values for ancestral spike.

5.2. Posterior prediction on the validation dataset

To assess the predictive capacity of the model fit, we plot the measured biomarker data for the validation model and $Y_{B, t_{i}}$, $Y_{P, t_{i}}$and $Y_{A, t_{i}}$ against the posterior predictive distributions of $X_{P}(t_{i})$, $X_{B}(t_{i})$, $X_{A}(t_{i}).$

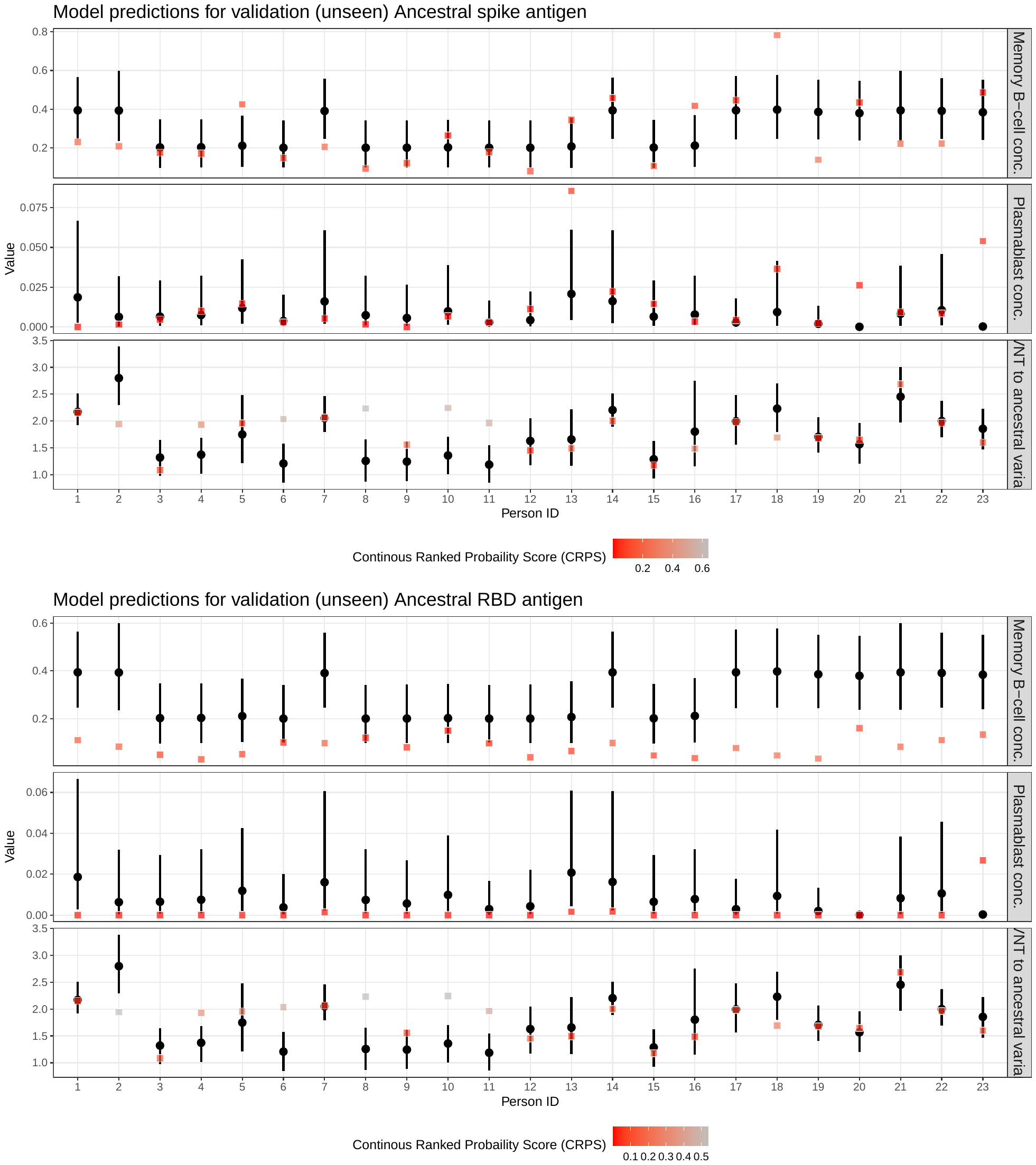

**Figure S5.** Comparison of the data and the model-predicted values for ancestral RBD.

Similarly, the individual-level fits for spike and RBD model on the validations data for all three biomarkers are shown in Figure x.

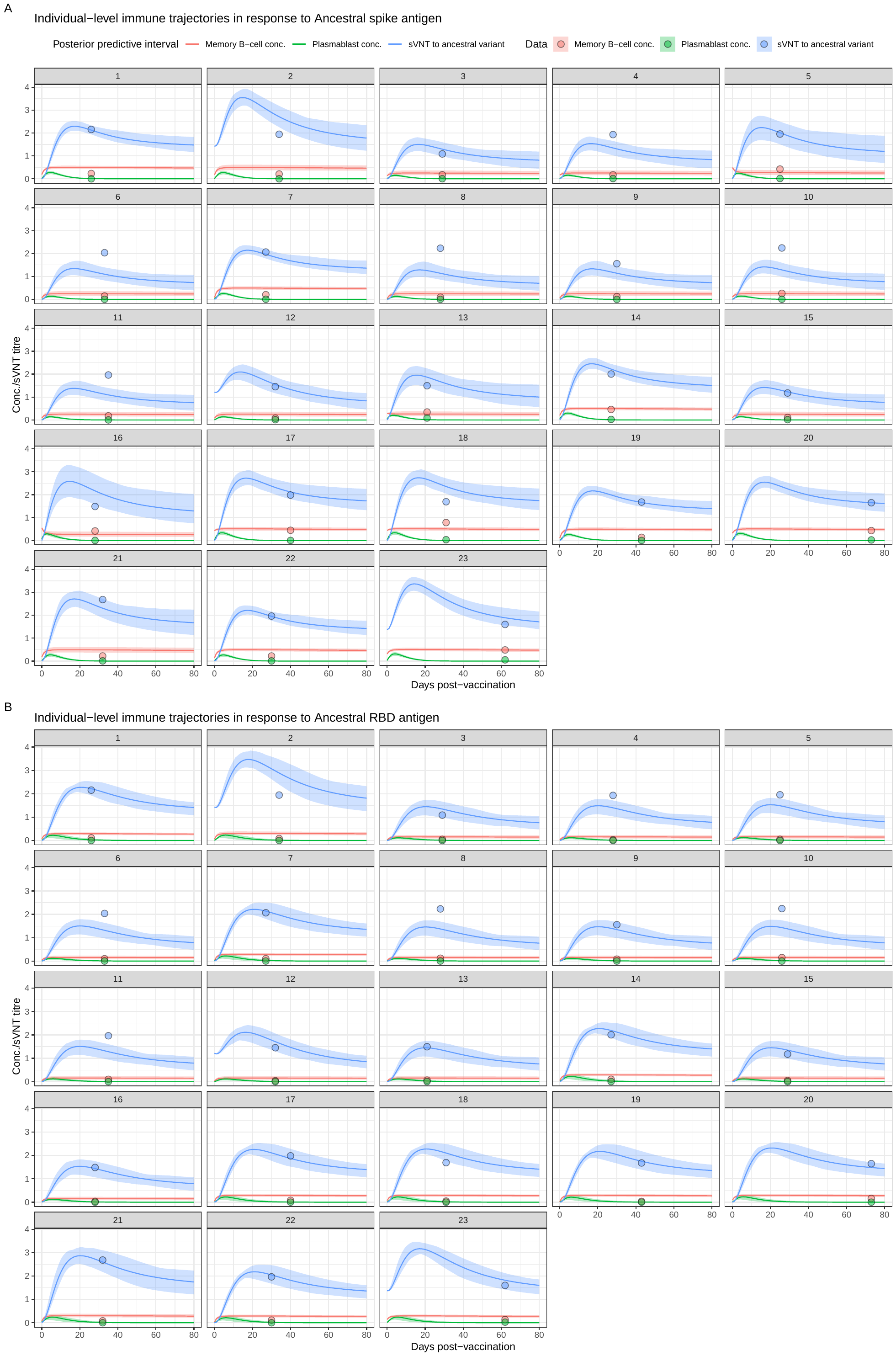

**Figure S6**. Individual-level trajectories comparing the data and the model-predicted values for ancestral RBD.

5.3. Posterior distributions of the fitted parameters.
     5.3.1. Spike model

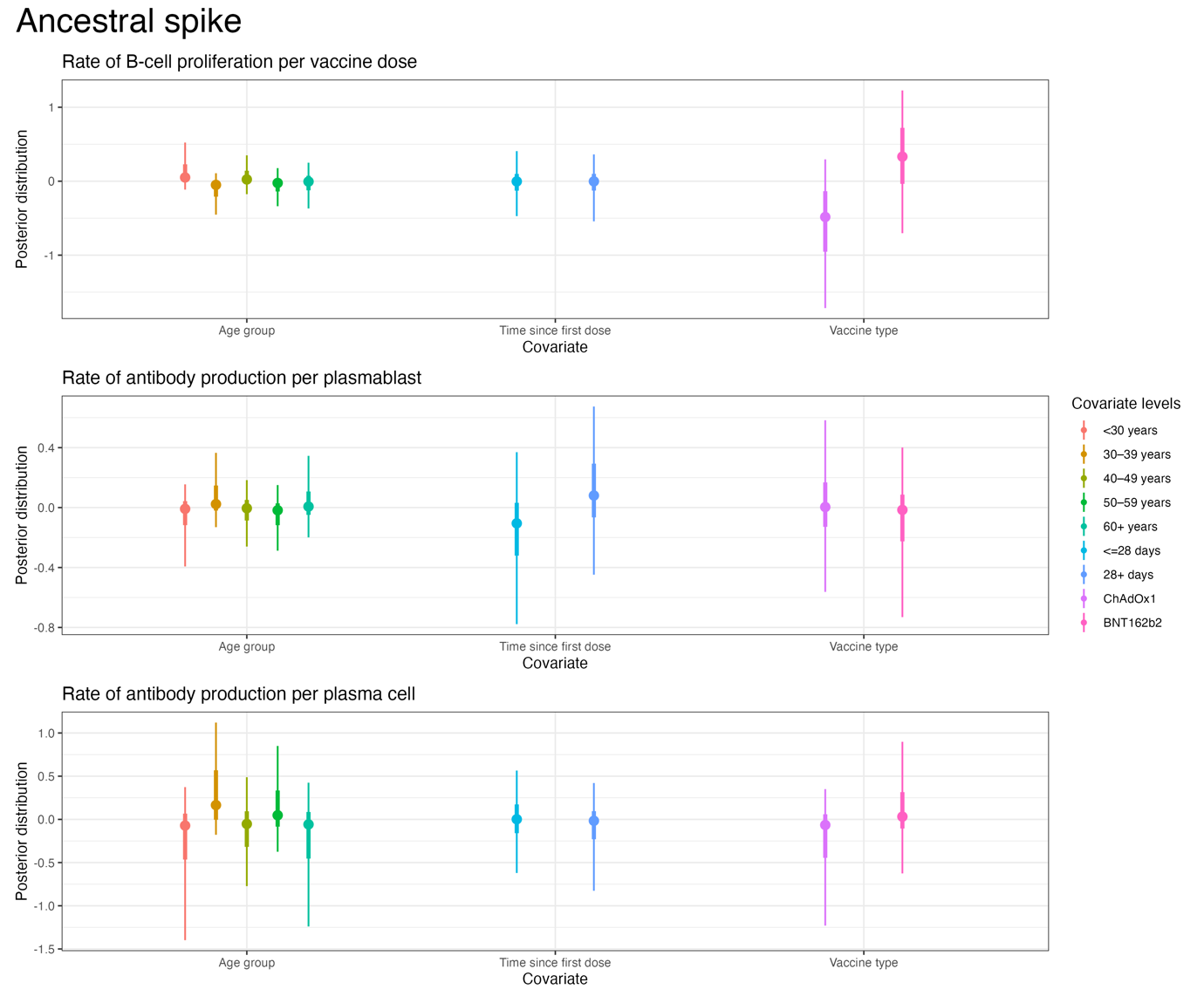

**Figure S7.** The posterior distribution for the hierarchical effects parameters for the spike fitting.

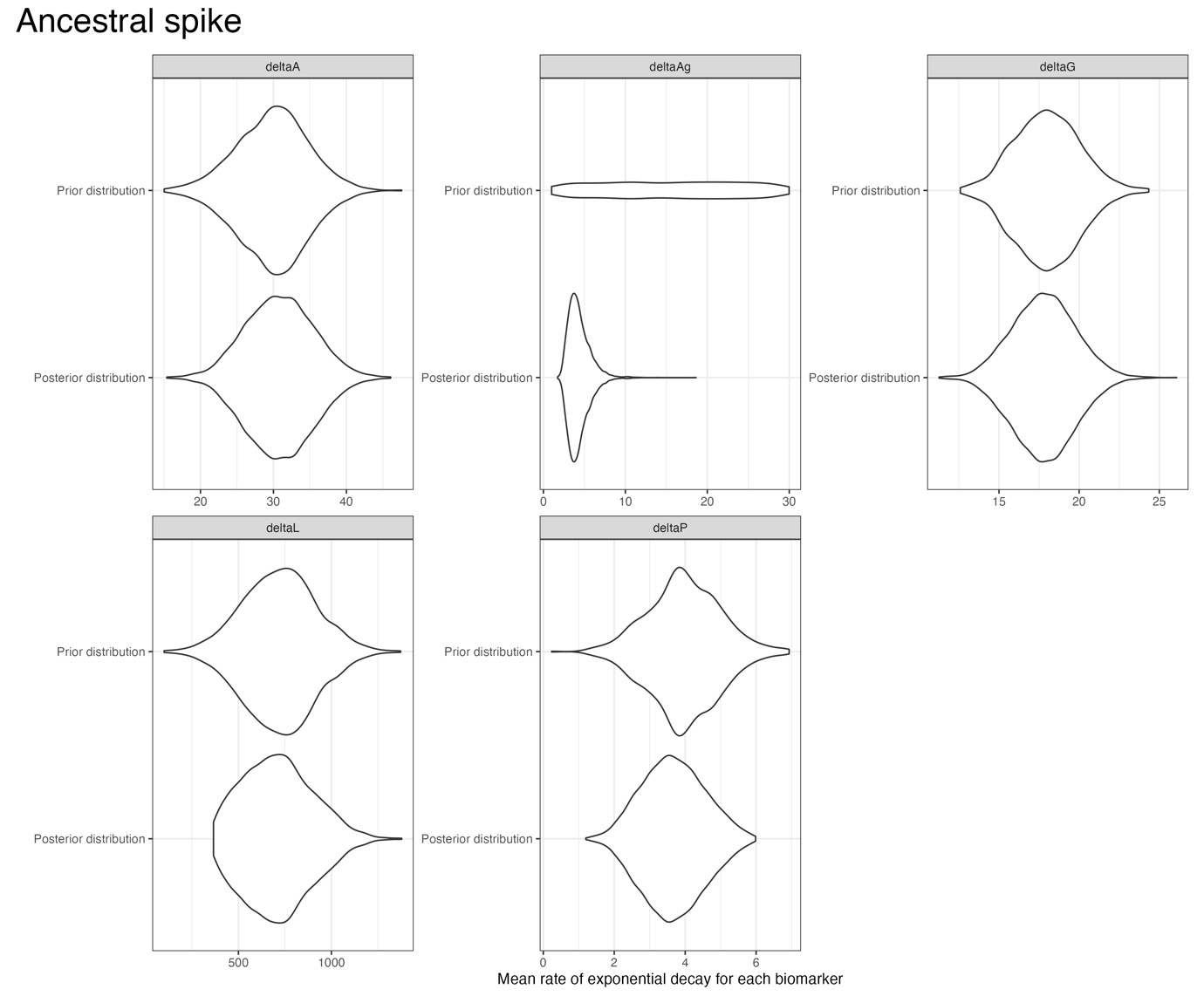

**Figure S8.** Posterior and prior distributions for the decay parameters in the spike fitting.

5.3.2. RBD model

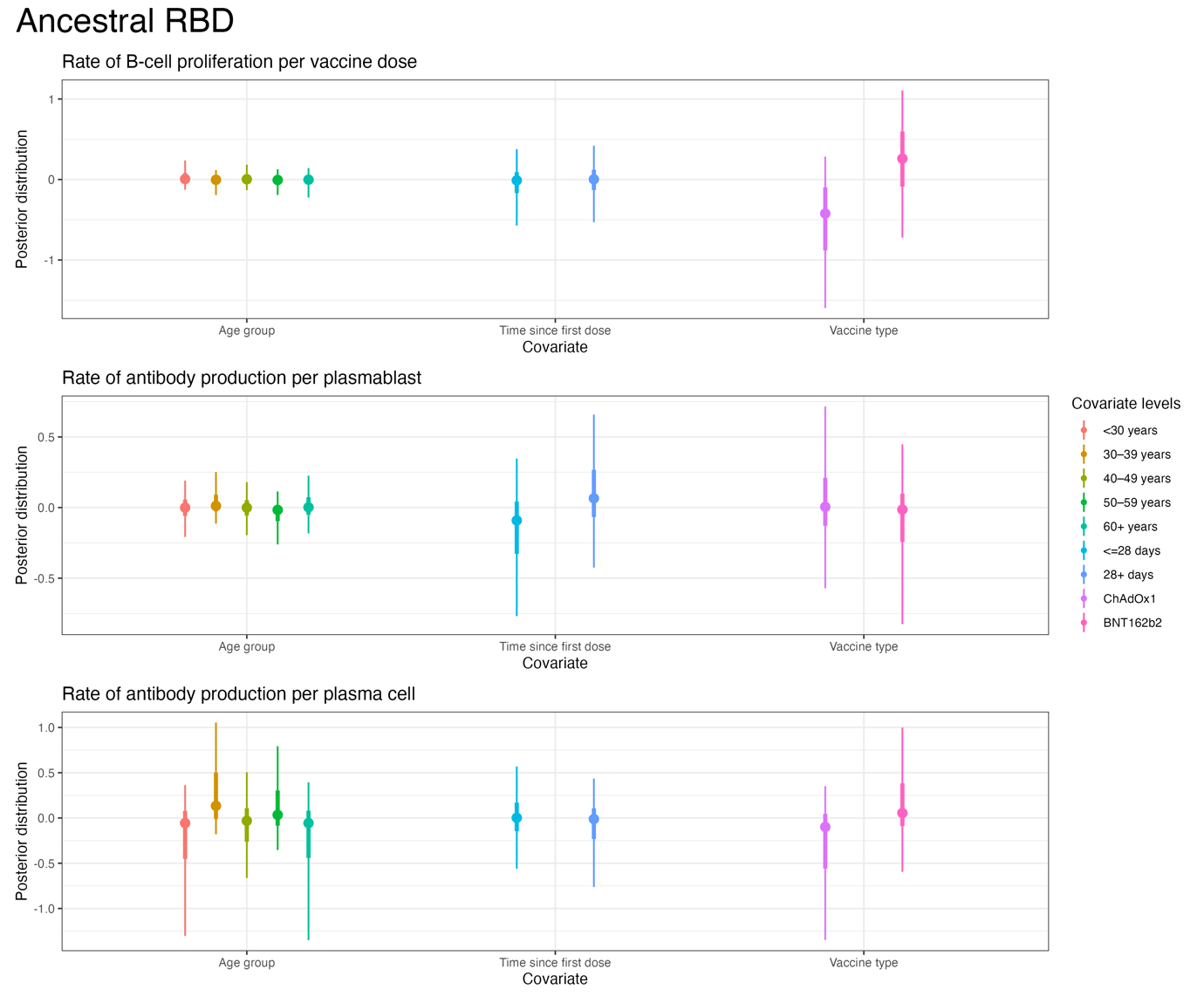

**Figure S9.** The posterior distribution for the hierarchical effects parameters for the RBD fitting.

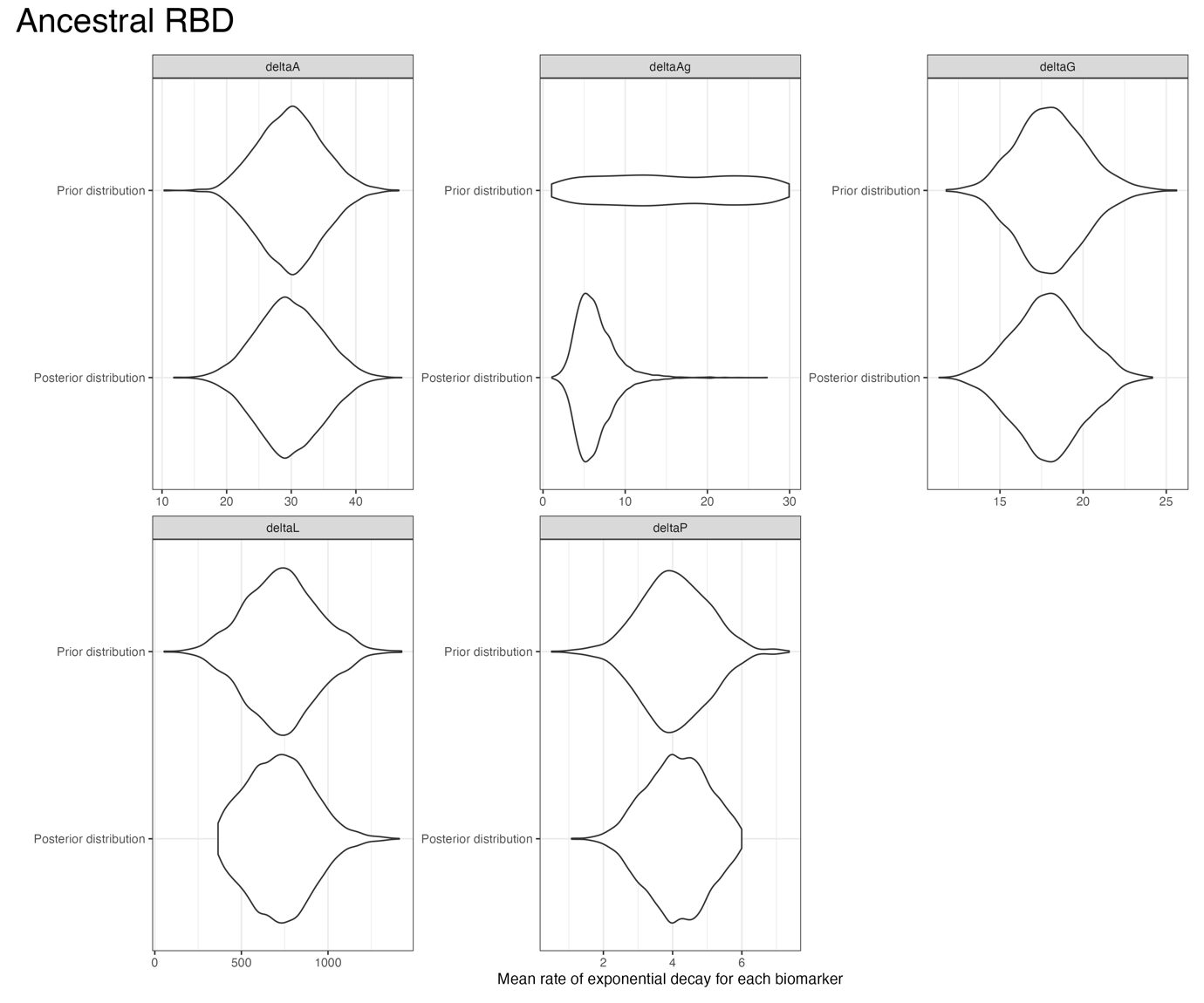

**Figure S10.** Posterior and prior distributions for the decay parameters in the RBD fitting.

5.4. Drivers of memory B cell proliferation and antibody production

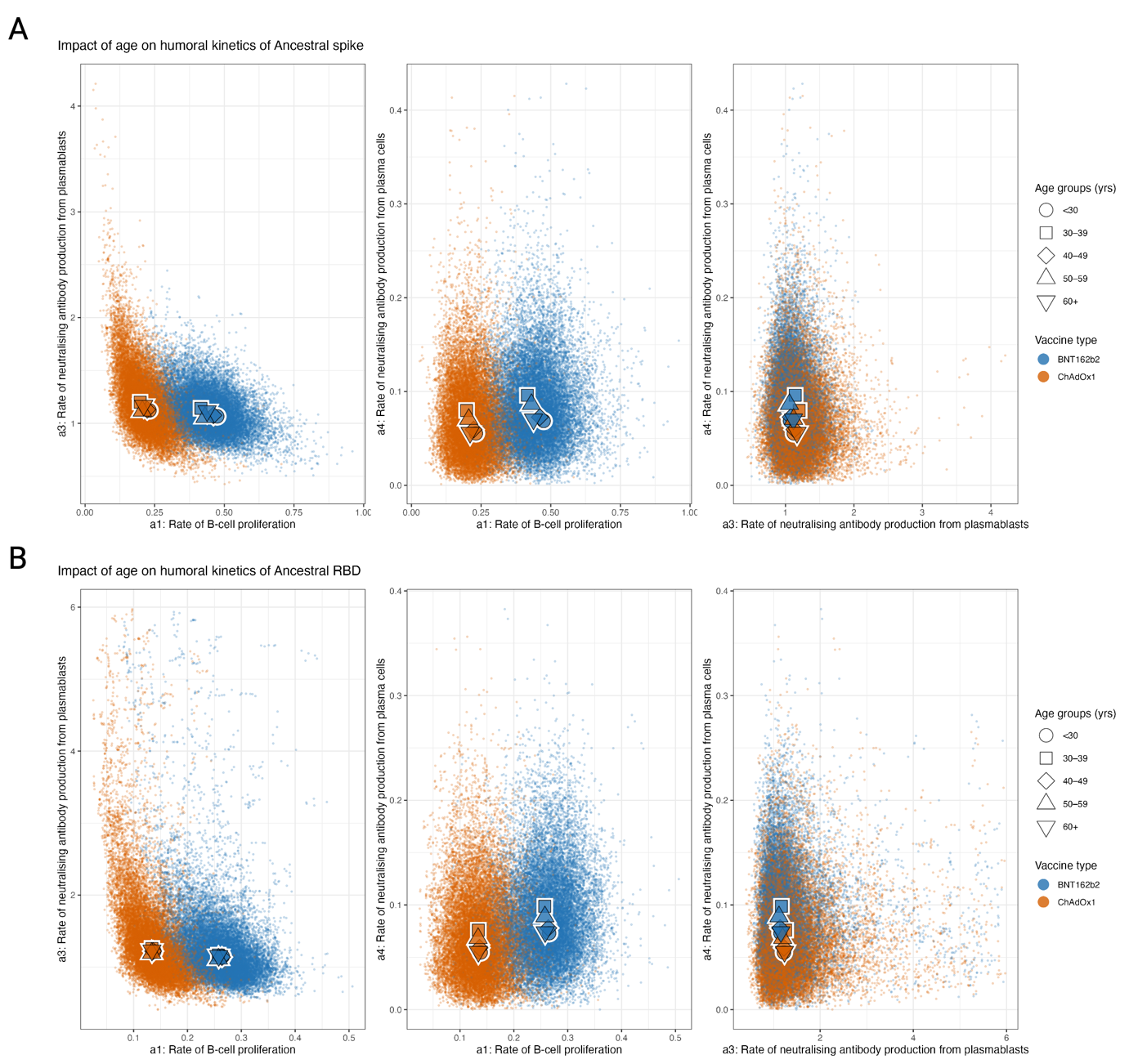

**Figure S11.   Posterior predictive distributions of the parameters driving the in-host dynamic model according to key covariates for ancestral spike and RBD stratified by age group** (A) Posterior predictive distributions for the impact of time since first dose on the rate of B cell proliferation (a_1_), the rate of neutralising antibody production from plasmablasts (a_3_), rate of neutralising antibody production from plasma cells (a_4_). (B) Posterior predictive distributions for the age on a_1_, a_3_, and a_4_.

5.5. Mechanistic predictions of antibody kinetics

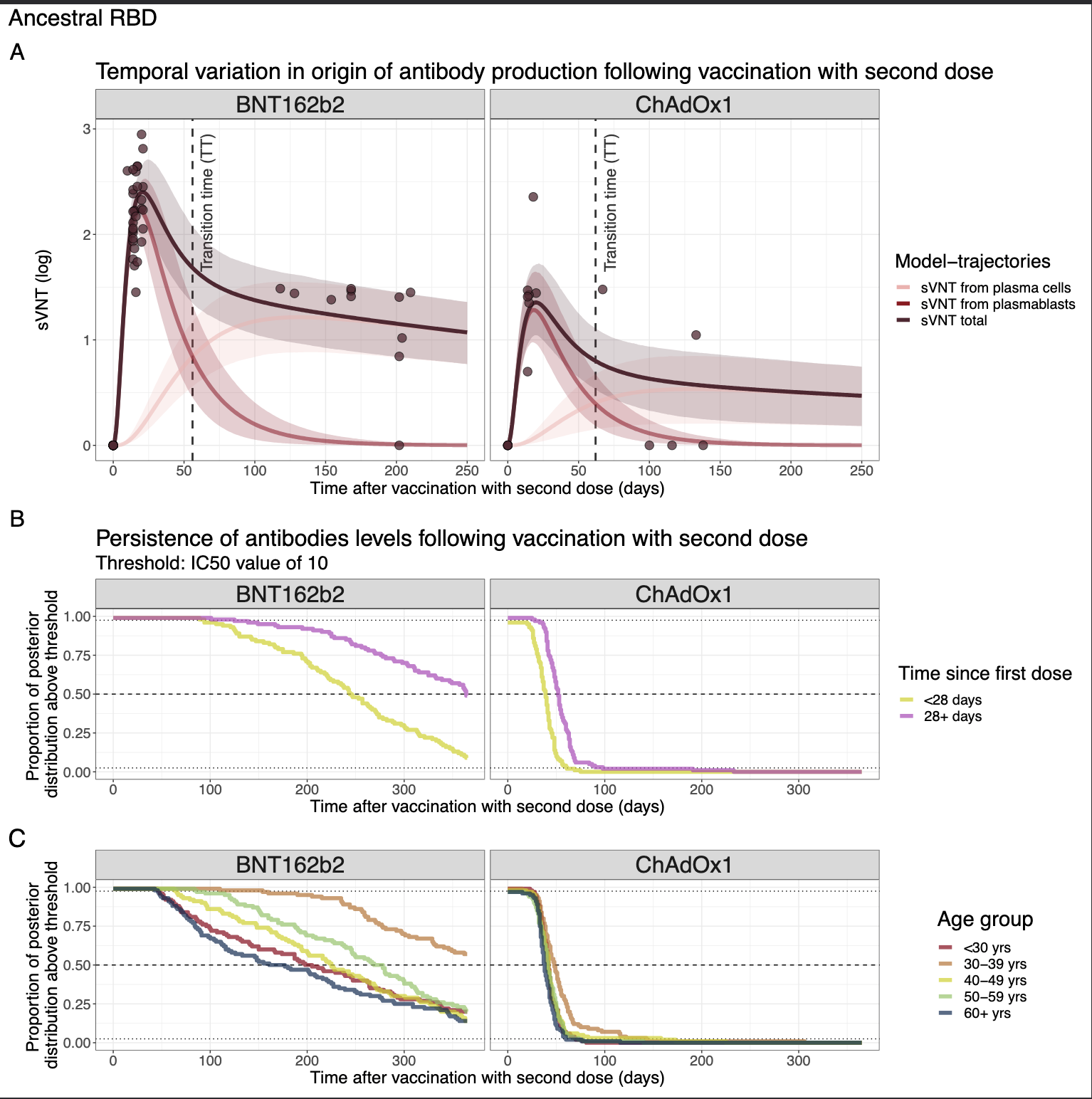

**Figure S12.** (A) Source of neutralising antibodies by days post-vaccination for ancestral RBD. Line and ribbons show the mean and 95% posterior predictive interval (PPI) and dots represent the sVNT from data. (B­–C) Complementary CDF of the marginal posterior distributions for the time antibody titres are above an IC50 threshold of 10 stratified by vaccinet type and B) time since last dose or C) age group.

**REFERENCES**

Flowers, Emily M., and Stephanie C. Eisenbarth. 2024. “In a Wild Germinal Center, What Determines Survival of the Fittest?” *Science Immunology* 9 (91): eadn7535.

Foss, Stian, Siri A. Sakya, Leire Aguinagalde, Marta Lustig, Jutamas Shaughnessy, Ana Rita Cruz, Lisette Scheepmaker, et al. 2024. “Human IgG Fc-Engineering for Enhanced Plasma Half-Life, Mucosal Distribution and Killing of Cancer Cells and Bacteria.” *Nature Communications* 15 (1): 2007.

Khodadadi, Laleh, Qingyu Cheng, Andreas Radbruch, and Falk Hiepe. 2019. “The Maintenance of Memory Plasma Cells.” *Frontiers in Immunology* 10 (April): 721.

Liu, Yi, Stephany Sánchez-Ovando, Louise Carolan, Leslie Dowson, Arseniy Khvorov, A. Jessica Hadiprodjo, Yeu Yang Tseng, et al. 2023. “Superior Immunogenicity of MRNA over Adenoviral Vectored COVID-19 Vaccines Reflects B Cell Dynamics Independent of Anti-Vector Immunity: Implications for Future Pandemic Vaccines.” *Vaccine* 41 (48): 7192–7200.

Russell, Timothy W., Hermaleigh Townsley, Sam Abbott, Joel Hellewell, Edward J. Carr, Lloyd Chapman, Rachael Pung, et al. 2023. “Within-Host SARS-CoV-2 Viral Kinetics Informed by Complex Life Course Exposures Reveals Different Intrinsic Properties of Omicron and Delta Variants.” *MedRxiv : The Preprint Server for Health Sciences*, May. https://doi.org/10.1101/2023.05.17.23290105.
